# Exome Array Analysis of 9,721 ischemic stroke cases from the SiGN Consortium

**DOI:** 10.1101/2022.11.09.22281914

**Authors:** Huichun Xu, Kevin Nguyen, Brady Gaynor, Hua Ling, Wei Zhao, Patrick F. McArdle, Timothy O’Connor, O Colin Stine, Kathleen A. Ryan, Megan Lynch, Jennifer A. Smith, Jessica D. Faul, Yao Hu, Jeffrey W. Haessler, Myriam Fornage, Charles Kooperberg, the Trans-Omics for Precision Medicine (TOPMed) Stroke Working Group, James A. Perry, Charles C. Hong, John W. Cole, Elizabeth Pugh, Kimberly Doheny, Sharon L.R. Kardia, David R. Weir, Steven J. Kittner, Braxton D. Mitchell, the SiGN Consortium

## Abstract

Recent studies have identified > 40 genetic variants robustly associated with ischemic stroke, most identified through genome wide association studies and primarily marking common variants in non-coding regions presumed to have regulatory roles on gene and protein expression. To evaluate the contribution of coding variants, which are mostly rare, to the etiology of ischemic stroke, we performed an exome array analysis of 9,721 ischemic stroke cases with mean age of onset at 67.1 years from the SiGN Consortium, and 12,345 subjects with no history of stroke (mean age 67.0 years) from the Health Retirement Study and SiGN consortium. Both cohorts included people with diverse ancestries. Genotyping for both SiGN and HRS was performed using similar array content at the Center for Inherited Disease Research (CIDR), albeit as two separate studies. Following extensive SNP- and sample-level quality control, a total of 106,101 SNPs from the exome content was used for exome association analysis. We identified 15 coding variants significantly associated with all ischemic stroke at array-wide threshold for statistical significance (i.e., p < 3.6 × 10^−7^) that also showed good genotyping quality, including two common SNPs in *ABO* that have previously been associated with stroke. Twelve of the remaining 13 variants were extremely rare in European Caucasians (MAF<0.1%) and the associations were driven by substantially higher allele frequencies in African American cases than in African American controls. A variant in *PRIM2*, rs199585353, was present exclusively in the stroke cases of European Caucasians while absent in all other samples from our data. There was no evidence for replication of these associations in either TOPMed Stroke samples (n = 5613 cases) or UK Biobank (n = 5,874 stroke cases), although power to replicate was very low given the low allele frequencies of the associated variants. In conclusion our analyses revealed 13 novel associations, but the low allele counts of associated variants and difficulty in acquiring large, well-powered replication highlight the challenges of rare variant association analysis, especially using array-based genotyping technologies.

## Introduction

Stroke is the second leading cause of disability and death worldwide, accounting for over 6 million deaths in 2019.^1^ The etiology of ischemic stroke, the predominant form of stroke, is multifactorial and includes both genetic and nongenetic causes. Genome-wide association studies (GWAS) have identified 89 stroke-associated loci to date ^2; 3^, although these loci account for only a very small proportion of stroke heritability. A major limitation of current genome-wide approaches, which rely predominantly on genotyping arrays, is that they typically interrogate only common variation throughout the genome (eg, SNPs with minor allele frequency > 1-5%) and generally do not cover the coding regions of the genome. Protein-coding variants are generally rare and are poorly captured by conventional GWAS arrays. Identifying the contribution of protein coding variation to stroke etiology is important. Even if exonic variation accounts for only a small proportion of stroke burden, identification of variants in novel genes may provide new insights into stroke biology.

The potential contribution of rare protein-coding variation to the etiology of ischemic stroke has not been systematically studied. Several small pilot exome-array association studies have been published based on relatively small numbers of subjects.^4^ In 2015, Auer et al, published an exome-wide association analysis based on 365 ischemic stroke cases with small- and large-vessel subtypes (plus additional controls) who underwent whole exome sequencing through the NHLBI Exome Sequencing Project.^5^ This study identified two protein-coding variants associated at exome-wide levels of significance, one a common variant (in *PDE4DIP*), and a second a rare variant (in *ACOT4*), although neither association has been replicated in subsequent studies. Using whole genome sequencing data from the TOPMed Consortium, Hu et al, recently performed a genome-wide analysis of 5,616 ischemic stroke cases and > 27,000 controls, from which they identified 2 variants significantly associated with IS and a 3^rd^ variant associated with LAS (n = 352 cases). The lead variants at all loci were low-frequency and more common in non-European populations. None of the variants were exonic, and none of these associations have been replicated in independent data sets, although the minor allele frequencies of these variants were low and the power to replicate limited.

To expand these efforts, we have performed an exome-wide array analysis of 9,721 stroke cases from the SiGN Network and 12,345 controls to evaluate the impact of rare coding variation on stroke risk.

## Methods

### Samples and genotyping

This study includes 9,721 ischemic stroke cases from the Stroke Genetics Network (SiGN) (dbGap Accession phs000615.v1.p1) and 12,345 non-stroke controls (1,303 from SiGN and 11,042 from the Health and Retirement Study. SiGN is an international collaboration of 31 studies across North America, Europe, and Australia to identify genetic determinants of ischemic stroke.^6^ The analysis presented in this manuscript includes subjects (mostly stroke cases) recruited from multiple sites in the United States and Europe (UK, Poland, Belgium, Spain, Austria, and Sweden). The HRS is a representative sample of people in the U.S. over the age of 50 residing in households with an oversample of African-American and Hispanic populations.^7^ HRS exome chip data is available with an approved HRS Restricted Data Agreement (RDA). Access information can be found at https://hrs.isr.umich.edu/data-products/genetic-data/products. Although exome array genotyping was successfully performed on 15,561 HRS subjects, only the subset of 11,042 HRS subjects who had no stroke history and also had genome-wide array data available were included in this study so that alignment of the genome-wide genotyping data could be used for estimation of ancestry.

Genotyping for both studies was performed at the Center for Inherited Disease Research (CIDR), SiGN cases on the Illumina HumanOmni5Exome-4v1_A array and HRS controls on the Illumina HumanExome-12v1-1 array. Both studies used calling algorithms implemented in GenomeStudio version 2011.1, Genotyping Module 1.9.4, and GenTrain version 1.0.

### Genotype quality control

A challenging feature of our study design is the use of cases and controls genotyped on slightly different arrays and at different times. We have previously performed a high quality GWAS of stroke in SIGN using external controls,^6^ which focused on common variants (MAF > 1%) as opposed to exome content enriched for low frequency variants. Rare variants are more challenging to call using array technologies because arrays rely on clustering due to genotype intensity to make genotype calls. To minimize the potential for bias arising from differential quality of genotyping calling between the two genotyping platforms, we therefore implemented a very stringent quality control procedure to identify poor quality SNPs and SNPs showing evidence for differential genotyping calling between the two arrays. In Stage 1 of our genotype quality control procedure, performed prior to association analysis, we utilized a large set of variant filters to identify and exclude SNPs of poor quality or differential quality between the two arrays. All remaining SNPs then underwent association analysis, after which we performed a Stage 2 quality control assessment that consisted of manual inspection of the genotype intensity plots of all associated SNPs from both the SiGN and HRS arrays to further exclude SNPs showing evidence of poor clustering on one or both genotype intensity plots. Manual inspection of genotype intensity plots for all SNPs prior to analysis was considered too labor-consuming and not feasible.

### Population structure analysis using Admixture and PCA

GWAS array genotypes from SIGN (Illumina 5MplusExome array) and HRS (Illumina Human Omni-2.5 Quad beadchip) were used for genetic ancestry analysis following genotype data cleaning as previously. ^6^ Only directly genotyped autosomal variants with minor allele frequency (MAF) > 5% were used. The variants were further pruned to keep independent variants not in linkage disequilibrium (LD). Principal component (PC) analysis of genotypes was carried out in Plink on unrelated samples and then the related samples were projected to the established PC space. Up to 10 PCs were included as covariates for association testing. Cases and controls had comparable distribution on the PC space. Particularly for “AFR” samples, there was no statistically significant difference in EUR or AFR component between cases and controls.

We additionally estimated the percentage of genetic ancestry (Europe, Africa, Native America, Eastern/South Asia) in individuals using the ADMIXTURE software program^8^ and the Human Genome Diversity Project (HGDP) reference genomes.^9^ Samples estimated as having genetic ancestry of (European + Central Asia) > 70% were classified as “EUR”, and samples estimated as African ancestry > 50% <u>and</u> Native American ancestry < 5% <u>and</u> Asian ancestry < 5% were classified as “AFR”. The remaining samples (mostly Latinx) were classified as “Other.” This classification was used to facilitate genotype cleaning and filtering as applied to a particular genetic ancestry, e.g comparing MAF between EUR samples from SiGN and EUR samples from HRS. Only “EUR” and “AFR” samples were used for this purpose.

### Association analysis

We performed association analysis using the mixed model as implemented in SAIGE.^10^ Genetic relationship matrix was modeled as a random effect. Covariates in the logistic regression model included sex and the first 10 principal components to account for ancestry. Power calculations indicated that our sample provided 80% power to detect odds ratios ranging from 1.09 to 1.20 for genetic variants with minor allele frequencies (MAF) = 0.5% and 1%, respectively, at the exome-wide threshold for significance, i.e., 4.7 × 10^−7^ for 106,101 variants.

### Replication

We sought to replicate associations in the TOPMed Consortium, through a look-up from the TOPMed Stroke Working Group. TOPMed Stroke included 5,613 ischemic stroke cases and 27,106 controls who underwent whole genome sequencing.^11^ Among the stroke cases were 4,305 cases of European ancestry and 884 cases of African ancestry. We additionally attempted replication of associated variants with stroke in the UK Biobank. For these analyses, we extracted ischemic stroke cases using an ICD code algorithm previously published in the “Definitions of Stroke for UK Biobank Phase 1 Outcomes Adjudication.”^12^ Ischemic stroke was defined using ICD 10 codes 163.X (cerebral infarction) and 164.X1 (stroke not specified as haemorrhage or infarction) and analyses performed in 5,874 stroke cases and 117,442 controls (i.e., 20 controls/case). All data were downloaded from the UK Biobank Resource under Application Number 49852. We performed logistic regression in PLINK using age, sex and 5 principal components for ancestry.

## Results

### Study subject characteristics

Our analysis was based on 9,721 cases and 12,345 controls. SiGN cases were recruited from 22 sites across the U.S. and Europe (Table S1). SiGN cases (all sites) plus SiGN controls from Belgium and Poland were genotyped on the Illumina HumanOmni5Exome-4v1_A array and all HRS controls were genotyped on the Illumina HumanExome-12v1-1 array. Characteristics of study subjects are shown in Table 1. The mean age of stroke onset in cases was 67 years (range: 14-104 years). 81% of cases and 80% of controls were genetically defined as European ancestry and 11% of cases and controls were genetically defined as African ancestry. TOAST subtype classification was unavailable in 64% of all stroke cases.

**Table 1.** Characteristics of study subjects

|  | Cases | Controls |
| --- | --- | --- |
| N | 9,721 | 12,345 |
| Age (or Age of onset for cases) | 67.0 ± 14.0 | 57.0 ± 9.7 |
| Age range (yrs) | 14-104 | 17-94 |
| % Female | 4662 (73.4%) | 7335 (78.2%) |
| Self-reported ancestry |  |  |
| EUR | 7138 (73.4%) | 9659 (78.2%) |
| AFR | 1022 (10.5%) | 1358 (11.0%) |
| Hispanic | 893 (9.2%) | 1034 (8.4%) |
| Other | 666 (6.9%) | 0 |
| unspecified | 2 (0.2%) | 294 (2.4%) |
| Genetic ancestry (computed) |  |  |
| EUR | 7921 (81.5%) | 9911 (80.3%) |
| AFR | 1044 (10.7%) | 1353 (11.0%) |
| Other | 756 (7.8%) | 1081 (8.8%) |
| # subjects excluded in QC analysis, which were based on unrelated subjects (but included in association analysis) | 246 (at $\pi\text{-hat} > 0.1875$) | 0 |
| TOAST classification |  |  |
| CE | 2210 (10.0%) |  |
| LAA | 1297 (5.9%) |  |
| OTHER | 296 (1.34%) |  |
| SAO | 2218 (10.1%) |  |
| UNDETERMINED | 1831 (8.3%) |  |
| UNKNOWN | 14214 (64.4%) |  |

### Merging of variants from the SiGN and HRS arrays

A total of 4,278,837 GWAS plus exome content SNPs were released for analysis in SiGN and 228,088 exome array SNPs for HRS following completion of array-specific initial quality control procedures by the genotyping center (CIDR) and subsequent in-depth quality assurance/quality control (QA/QC) analysis by the genotyping analysis core at the University of Washington (SiGN) or the University of Michigan (HRS). The sample level and variant level filters recommended by the genotyping analysis cores were applied before data merging and the two-stage variant QC described below for current study. After removing variants that failed strand or allele alignment, there were a total of 198,811 overlapping SNPs between the SiGN 5MPlusExome array. We then removed 10,413 of these aligned SNPs because they were peripheral to relevant exome content, as they were included as ancestry-informative variants or for QC or method development purposes. We then conducted detailed quality control analyses on the remaining set of 188,398 SNPs merged from SiGN and HRS to identify SNPs whose genotyping quality potentially differed across platforms.

### Stage 1 variant quality control filtering

We implemented multiple quality control checks to exclude potentially problematic SNPs from analysis. Criteria for excluding SNPs included: (1) excessive deviation from HWE in either EUR or AFR controls, or extreme deviation in cases; (2) AT/GC SNPs with high allele frequency; (3) discordant genotype calls between samples genotyped on both platforms; (4) excessive differences in AF between controls genotyped in SIGN and HRS; (5) low minor allele count; (6) high genotype missingness calls; (7) possible under-calling of genotypes, especially in SiGN (8) non-autosomal SNPs, (9) SNPs marked in HRS as technical failures; (10) large AF differences between controls and gnomAD; and (11) SNPs with duplicate probes but discordant genotypes (Figure 1). A total of 82,297 (43.7%) of SNPs were excluded, although the bulk of these (n = 78,725) were excluded due to low MAC in both cases and controls. A detailed description of the variant exclusion criteria, including filters for used for each criterion, is provided in the Appendix, and the specific filtering thresholds used at each step in QC are provided in Table 2. For HRS, we applied the filters recommended in the Quality Control Report recommended by HRS Analysis team at the University of Michigan.

**Table 2:** SNP Quality control filters

|  |  |  |
| --- | --- | --- |
| **1. MERGE HRS + SiGN** |  | 198,811 SNPs |
| **2. Remove probes added by design (eg, ancestry-specific variants and SNPs added for genotyping QC, as previously annotated in CHARGE (see text))** |  | 188,398 SNPs |
|  |  | **Excluded # of SNPs\*** |
| **3. SNP QC filter** |  |  |
|  | SNPs with genotype AT or GC if AF is between 40-60% | 132 |
|  | extreme deviation from HWE | 3,552 |
| | In controls from HRS study: ($p < 1E-5$ (EUR), $1E-10$ *AFR in HRS | 162 |
| | in controls from SIGN dataset: $p < 1E-5$ (EUR) | 51 |
| | in stroke cases: ($p < 1E-20$ (EUR), $1E-10$ (AFR)) | 3,552* |
|  | discordant call > 0 among technical duplicated samples from 51 subjects genotyped on both platforms (29 HRS samples and 22 WUSTL samples) | 205 |
| | large allele frequency differences in EUR (EWAS $p < 1E-3$) between control samples genotyped in SiGN and HRS | 147 |
| **4. Missing rate** |  |  |
| | > 2.5% in SiGN or HRS for non-rare variants with MAF $\geq 1\%$, | 1,145 |
|  | > 0.8% in SiGN or HRS for rare variants with MAF < 1% | 43,289 |
| | Possibly undercalled in SiGN: MAC $\leq 3$ in SiGN (all ancestries), but > 20 in HRS EURs | 66 |
|  | Marked as technical failure in HRS upon additional manual review of Genome Studio clustering plots | 308 |
| | Allele freq differ significantly between HRS EUR samples and GNOMAD non-Finnish EUR ($p < 1E-5$ for rare alleles, $p < 1E-10$ for low freq and common alleles) | 1,618 |
| | duplicate probes on SIGN 5MplusExome array with discordant allele feqs (Fisher exact $p < .05$) | 144 |
|  | SNPs with low MAC (<10) | 78,725 |
|  | Non-autosomal SNPs | 3,478 |
|  | **Number of SNPs remaining** | **106,101** |
| **4. SNP-specific and study site specific masking for variants from SIGN samples:** |  |  |
| | variants showed substantial allele frequency differences between a particular site and the remaining samples ($P < 1E-3$ and AC>10) | 993 |
|  | variants showed substantial call rate differences between a particular site and the remaining samples (differential missingness p value <5E-7) | 12691 |
|  | **Number of SNPs remaining** | **106,101** |

\* excluded SNPs are not exclusive; eg.. 3,534 of these SNPs also excluded by other criteria

**Figure 1:**
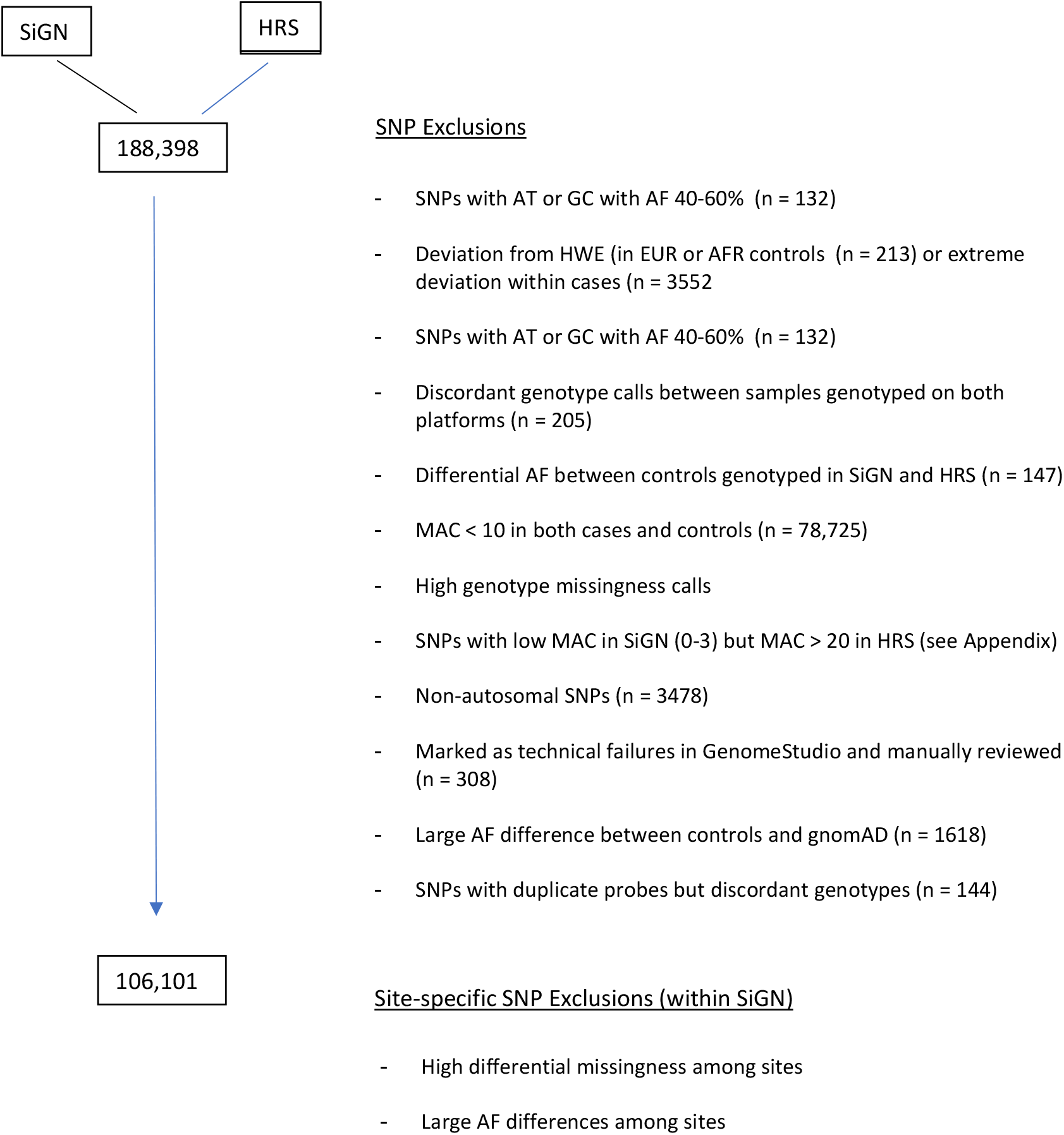
Stage 1 variant quality control pipeline. 188,398 SNPs merged between the SIGN and HRS arrays. 82,297 SNPs excluded, leaving 106,101 SNPs for analysis (see Table S1 for specific filtering thresholds at each QC step)

### Genetic association results

Following variant quality control filtering, we performed genetic association studies on 106,101 variants with minor allele count ≥ 10 in cases or controls. We identified 36 variants significantly associated with all ischemic stroke meeting array-wide threshold for statistical significance (i.e., p < 3.6 × 10^−7^). Upon manual review of these variants, we considered the SiGN or HRS genotype intensity plots for 21 of these SNPs to be of poor or questionable clustering quality, leaving 15 variants that we regarded as being robustly associated with stroke. Association results for these variants are shown for the overall meta-analysis in Table 3, and the genotype intensity plots for these 15 associated SNPs from both the SiGN and HRS arrays are provided in Supplemental Figure 1. Among the 15 associated variants were two common SNPs in *ABO*, rs507666 and rs635634 that are in near perfect linkage disequilibrium with each other (r^2^ = 0.99) and that have previously been associated with stroke.^2^ Twelve of the remaining 13 variants were extremely rare in European Caucasians (MAC ≤ 10 combined for each in cases and controls) and the associations were driven by substantially higher allele frequencies in African American cases than in African American controls (Table 4). Four of these variants (*TPTE* rs143510517, *MEP1A* rs62619974, *DDX31* rs142792732, and *PATL1* rs79336999) were associated with stroke at p < 1 × 10^−10^. A variant in *PRIM2*, rs199585353, was present exclusively in European Caucasians, in whom the minor allele frequency was 0.25% in cases (MAC = 40) and 0% in controls (p = 8.28 × 10^−8^).

**Table 3.** 15 SNPs associated with stroke on exome-wide analysis

| SNP | rs number | Gene | Function | AAchg | A2 | A1 | OR | p-value |
| --- | --- | --- | --- | --- | --- | --- | --- | --- |
| exm1345082 | rs192153785 | GH2 | missense | Q228P | T | G | 7.45 | 1.92E-08 |
| exm1501517 | rs140922537 | ZNF765 | missense | P270S | C | T | 6.99 | 5.63E-10 |
| exm1562153 | rs143510517 | TPTE | missense | R274W | G | A | 6.56 | 7.17E-12 |
| exm21949 | rs373898350 | NBPF1 | unknown |  | G | T | 6.73 | 3.56E-07 |
| exm365204 | rs141845742 | SPATA16 | stoploss | X570Q | A | G | 7.90 | 2.56E-07 |
| exm384695 | rs149905649 | DOK7 | missense | R92W | C | T | 0.16 | 3.67E-08 |
| exm552854 | rs62619974 | MEP1A | missense | K396R | A | G | 6.77 | 1.04E-11 |
| exm558342 | rs199585353 | PRIM2 | unknown |  | G | T | 5.51 | 8.20E-08 |
| exm615057 | rs375144101 | TRGC1 | stoploss | Ter174LysextTer17 | A | T | 7.35 | 4.46E-10 |
| exm791656 | rs142792732 | DDX31 | missense | E33K | C | T | 0.16 | 2.38E-17 |
| exm90767 | rs372423248 | SEC22B | unknown |  | A | C | 7.41 | 2.57E-08 |
| exm90783 | rs373433490 | SEC22B | unknown |  | C | T | 6.74 | 1.65E-10 |
| exm913753 | rs79336999 | PATL1 | missense | Y645C | T | C | 7.52 | 2.10E-13 |
| exmrs507666 | rs507666 | ABO |  |  | G | A | 1.17 | 4.79E-08 |
| exmrs635634 | rs635634 | ABO |  |  | C | T | 1.17 | 6.12E-08 |

**Table 4.** Comparison of MAF and allele counts between cases and controls, by ancestry for 15 EWA stroke-associated SNPs

| SNP | rs number | Gene | A2/A1 | Ancestry = ALL |  |  |  |  | EUROPEANS ONLY |  |  |  |  | AFRICAN AMERICANS ONLY |  |  |  |  |
| --- | --- | --- | --- | --- | --- | --- | --- | --- | --- | --- | --- | --- | --- | --- | --- | --- | --- | --- |
|  |  |  |  | CASES<br>(n = 9,721) |  | CONTROLS<br>(n = 12,345) |  | MAF<br>gnomad<br>(TOTAL) | CASES<br>(n = 7,138) |  | CONTROLS<br>(n = 9,659) |  | MAF<br>gnomad<br>(EUR non-FINNISH)<br>V3 | CASES<br>(n = 1,022) |  | CONTROLS<br>(n = 1,358) |  | MAF<br>gnomad<br>(AFR/AFR<br>AM) |
|  |  |  |  | MAF | AC | MAF | AC |  | MAF | AC | MAF | AC |  | MAF | AC | MAF | AC |  |
| exm1345082 | rs192153785 | GH2 | T/G | 0.21% | 40 | 0.00% | 0 | 0.15% | 0.01% | 2 | 0.00% | 0 | 0.00% | 1.58% | 33 | 0.00% | 0 | 1.44% |
| exm1501517 | rs140922537 | ZNF765 | C/T | 0.25% | 49 | 0.01% | 2 | 0.25% | 0.03% | 5 | 0.00% | 0 | 0.01% | 1.92% | 40 | 0.04% | 1 | 2.43% |
| exm1562153 | rs143510517 | TPTE | G/A | 0.36% | 69 | 0.01% | 2 | 0.07% | 0.04% | 6 | 0.00% | 0 | 0.00% | 2.55% | 53 | 0.04% | 1 | 0.77% |
| exm21949 | rs373898350 | NBPF1 | G/T | 0.18% | 35 | 0.00% | 1 | 0.00% | 0.01% | 1 | 0.00% | 0 | 0.00% | 1.35% | 28 | 0.04% | 1 | 0.03% |
| exm365204 | rs141845742 | SPATA16 | A/G | 0.16% | 31 | 0.00% | 0 | 0.12% | 0.00% | 0 | 0.00% | 0 | 0.00% | 1.29% | 27 | 0.00% | 0 | 1.17% |
| exm384695 | rs149905649 | DOK7 | C/T | 0.00% | 0 | 0.22% | 54 | 0.22% | 0.00% | 0 | 0.06% | 10 | 0.09% | 0.00% | 0 | 1.45% | 39 | 1.66% |
| exm552854 | rs62619974 | MEP1A | A/G | 0.34% | 67 | 0.00% | 1 | 0.36% | 0.01% | 1 | 0.00% | 0 | 0.01% | 2.83% | 59 | 0.00% | 0 | 3.08% |
| exm558342 | rs199585353 | PRIM2 | G/T | 0.21% | 40 | 0.02% | 6 | 0.02% | 0.25% | 40 | 0.00% | 0 | 0.04% | 0.00% | 0 | 0.00% | 0 | 0.01% |
| exm615057 | rs375144101 | TRGC1 | A/T | 0.26% | 50 | 0.00% | 1 | 0.13% | 0.04% | 6 | 0.00% | 0 | 0.01% | 1.63% | 34 | 0.04% | 1 | 1.27% |
| exm791656 | rs142792732 | DDX31 | C/T | 0.00% | 0 | 0.58% | 143 | 0.37% | 0.00% | 0 | 0.03% | 5 | 0.00% | 0.00% | 0 | 4.77% | 128 | 4.11% |
| exm90767 | rs372423248 | SEC22B | A/C | 0.21% | 40 | 0.00% | 0 | 0.03% | 0.01% | 1 | 0.00% | 0 | 0.00% | 1.58% | 33 | 0.00% | 0 | 0.28% |
| exm90783 | rs373433490 | SEC22B | C/T | 0.27% | 53 | 0.01% | 3 | 0.01% | 0.02% | 3 | 0.00% | 0 | 0.00% | 2.11% | 44 | 0.12% | 3 | 0.11% |
| exm913753 | rs79336999 | PATL1 | T/C | 0.37% | 71 | 0.00% | 0 | 0.22% | 0.00% | 0 | 0.00% | 0 | 0.00% | 3.21% | 67 | 0.00% | 0 | 2.41% |
| exm-rs507666 | rs507666 | ABO | G/A | 20.80% | 3589 | 17.99% | 4442 | 17.23% | 22.93% | 3207 | 19.14% | 3297 | 19.85% | 9.95% | 183 | 10.43% | 281 | 10.79% |
| exm-rs635634 | rs635634 | ABO | C/T | 20.78% | 3586 | 18.00% | 4443 | 17.17% | 22.94% | 3209 | 19.13% | 3296 | 19.79% | 9.68% | 178 | 10.50% | 283 | 10.75% |

Of the 13 non-ABO variants, all have MAF < 0.1% in European ancestry populations and ≤ 4% in African ancestry populations as indicated in gnomAD. Seven of the 13 are annotated through ANNOVAR using RefSeq gene annotation ^13^ as missense variants, 2 as stoploss variants, and the remaining 4 as function unknown. One SNP rs149905649 in gene DOK7 was included in ClinVar but was annotated as “benign/likely benign”. None of the remaining SNPs were included in ClinVar.

### Replication

We sought to replicate the associations of these 15 variants in TOPMed Stroke and UKB. In TOPMed Stroke, 12 of the 15 variants were polymorphic in TOPMed, none of which showed evidence for association (Table S2). Associations of the two ABO variants were directionally consistent with the discovery analysis in both the European and African ancestry populations. Among European ancestry cases and controls combined, only 2 of the remaining 13 (non-ABO) variants had minor allele counts > 8; among African ancestry cases alone, 8 of these 13 variants had minor allele counts ≥ 14 (range 14-68).

In the UKB, we attempted to replicate only the *PRIM2* rs199585353 and *DOK7* rs149905649 associations observed in the European ancestry group as the number of African ancestry individuals in UKB with stroke was small (n = 101). The imputation quality of *PRIM2* rs199585353 did not meet our info score quality control threshold of 0.7 (info = 0.51) and was therefore not analyzed. The MAF for *DOK7* rs149905649 was 0.00051 in cases (6/(5874*2)) and 0.00068 in controls (159/(117439*2)); p = 0.46).

## Discussion

We combined exome array genotypes from a large collection of stroke cases with exome array data from publicly available controls to carry out a large-scale association study to identify rare protein-coding variants associated with ischemic stroke. With 9,721 well-phenotyped stroke cases, this study represents the largest effort to date to identify rare protein-coding variants associated with ischemic stroke at an exome-wide level. Although we identified 13 rare variants meeting exome-wide thresholds for association, none replicated in the 2 replication datasets. Of the 13 rare variants none have known biology function to be compelling candidate genes responsible for the pathogenesis of ischemic stroke; 12 of the 13 associations were driven by allele frequency differences in the African American population, in whom the sample sizes in both the Discovery and Replication data sets were much smaller. We also identified two common variants (MAF ∼ 0.20 in European ancestry) in *ABO* that are in high linkage disequilibrium with each other and that have been previously associated with ischemic stroke.^2^

Our results lend further support for advocating the inclusion of diverse populations in genomic studies. The sample sizes of African American (AA) samples were small in our discovery data (1044 AA cases). However, most of the identified exome variants in our EWAS analysis (12 out of 13 variants) reached statistical significance due to their presence or relatively higher minor allele frequencies in African American (MAF 0.03%∼4.11%), while being absent in Europeans. Nonetheless, at n=∼1,000 of cases, the power was too low for a reliable discovery and replication, especially given the genetic diversity among African American across study cohorts. It is thus not surprising that these AA-specific coding signals did not replicate in the TOPMed cohort, in whom the AA sample size was also limited. Our efforts highlight the need to expand genomics research in non-European populations.

Up to 81.5% of our discovery samples were of European ancestry genetically. Yet, we identified only 2 exome-wide associated loci driven by European samples, one previously identified at *ABO* and the second a novel locus in *PRIM2*, that was not replicated in either TOPMed or UKB. There are several possible reasons for our failure to identify more robust associations in exomes of Europeans. First, the coverage of the exome in our analysis was low due to our stringent filtering criteria and some ‘causally’ variants may have failed QC and not been tested. In fact, only 46.5% of the variants of the exome array content were actually tested. Whole exome sequencing would have provided much greater coverage across genomes. Second, the overall EWAS power to detect and replicate ‘significant’ associations for rare variants in our dataset, even with 9,721 cases in the Discovery set, was low. Although the two *ABO* variants are very common with MAF up to 20% in European, the variant in *PRIM2* only has MAF 0.04% in GNOMAD European samples. Third, it is also possible that rare protein-coding variants do not play a large role in the etiology of ischemic stroke.

Our study highlights a major challenge in accruing large sample sizes for rare variant analyses. The problem of accruing large samples sizes for analysis of common variants has been addressed successfully by combining study-specific genome-wide association results through meta-analysis. Within contributing studies, established protocols are typically used that include a thorough assessment of genotype intensity clusters for evaluation of cluster separation and genotype calls. Such assessments rely on sufficient numbers of each genotype to establish genotype separation boundaries. If sufficient numbers of each genotype are not available, as when the minor allele count is very small, establishing the genotype cluster separation boundaries can be difficult, making the genotype call unreliable. This problem is magnified if cases and controls are genotyped in different labs or different batches, because any ‘batch’ effect will mimic a difference between cases and controls. We attempted to address this problem by implementing a very stringent set of quality control measures that considered both within study (SiGN and HRS) as well as between study measures. We further attempted to minimize the possibility of identifying false results by manually inspecting the genotype intensity clusters of all variants we reported to be exome-wide associated with stroke. But the cost of our implementing this stringent procedure was removing a large proportion (46.6%) of variants from analysis. The rigid variant QC procedures coupled with the necessity of manually inspecting all genotype intensity plots for all ‘reportable’ associations also makes bin-based analyses much less attractive since many bins will be incompletely covered due to variant filtering and all variants would require the laborious task of evaluation of genotype intensity plots.

Future efforts to identify rare protein-coding variants associated with stroke would be wise to pay heed to these lessons by using studies that rely either on non-array genotyping technologies, such as sequencing, for variant detection, or to employ very large samples for array-based studies in which cases and controls are genotyped together with careful effort made to minimize batch effects. For example, a recently published study from the TOPMed Stroke Working Group was based on whole genome sequencing (WGS), although this study included only 5,616 ischemic stroke cases and 27,116 non-stroke controls.^11^ This WGS study identified five novel variants associated with stroke. However, only 2 of these variants were present in SiGN and neither provided evidence for replication.

Our study includes other limitations. Even with 9,721 stroke cases, our sample is powered only to detect those rare variants having relatively large effect sizes. Stroke subtype information was available for only 35.6% of our cases, even further limiting power for stroke subtype-specific analyses.

In summary, we have conducted the largest effort to date to identify rare protein-coding variants associated with ischemic stroke at an exome-wide level. We identified 13 rare stroke-associated variants as well as one additional association with 2 common variants at a previously known locus in ABO. Our study highlights the multiple challenges in using publicly available controls for large-scale rare variant array-based studies and the importance of expanding the inclusion of diverse non-European samples in the genetic study of ischemic stroke.

## Data Availability

All results produced in the present study are available upon reasonable request to the authors.

## References

1. World Health Organization. (2019). The top 10 causes of death. In. (

2. Malik, R., Chauhan, G., Traylor, M., Sargurupremraj, M., Okada, Y., Mishra, A., Rutten-Jacobs, L., Giese, A.K., van der Laan, S.W., Gretarsdottir, S., et al. (2018). Multiancestry genome-wide association study of 520,000 subjects identifies 32 loci associated with stroke and stroke subtypes. Nat Genet.

3. Mishra, A., Malik, R., Hachiya, T., Jürgenson, T., Namba, S., Posner, D.C., Kamanu, F.K., Koido, M., Le Grand, Q., Shi, M., et al. (2022). Stroke genetics informs drug discovery and risk prediction across ancestries. Nature 611, 115–123.

4. Jaworek, T., Ryan, K.A., Gaynor, B.J., McArdle, P.F., Stine, O.C. TD O.C., Lopez, H., Aparicio, H.J., Gao, Y., Lin, X., et al. (2020). Exome array analysis of early-onset ischemic stroke. Stroke 51, 3356–3360.

5. Auer, P.L., Nalls, M., Meschia, J.F., Worrall, B.B., Longstreth, W.T., Jr., Seshadri, S., Kooperberg, C., Burger, K.M., Carlson, C.S., Carty, C.L., et al. (2015). Rare and coding region genetic variants associated with risk of ischemic stroke: The NHLBI Exome Sequence Project. JAMA Neurol 72, 781–788.

6. NINDS Stroke Genetics Network, and International Stroke Genetics Consortium. (2016). Loci associated with ischaemic stroke and its subtypes (SiGN): a genome-wide association study. Lancet Neurol 15, 174–184.

7. Sonnega, A., Faul, J.D., Ofstedal, M.B., Langa, K.M., Phillips, J.W., and Weir, D.R. (2014). Cohort Profile: the Health and Retirement Study (HRS). Int J Epidemiol 43, 576–585.

8. Alexander, D.H., Novembre, J., and Lange, K. (2009). Fast model-based estimation of ancestry in unrelated individuals. Genome Res 19, 1655–1664.

9. Li, J.Z., Absher, D.M., Tang, H., Southwick, A.M., Casto, A.M., Ramachandran, S., Cann, H.M., Barsh, G.S., Feldman, M., Cavalli-Sforza, L.L., et al. (2008). Worldwide human relationships inferred from genome-wide patterns of variation. Science 319, 1100–1104.

10. Zhou, W., Nielsen, J.B., Fritsche, L.G., Dey, R., Gabrielsen, M.E., Wolford, B.N., LeFaive, J., VandeHaar, P., Gagliano, S.A., Gifford, A., et al. (2018). Efficiently controlling for case-control imbalance and sample relatedness in large-scale genetic association studies. Nat Genet 50, 1335–1341.

11. Hu, Y., Haessler, J.W., Manansala, R., Wiggins, K.L., Moscati, A., Beiser, A., Heard-Costa, N.L., Sarnowski, C., Raffield, L.M., Chung, J., et al. (2022). Whole-genome sequencing association analyses of stroke and its subtypes in ancestrally diverse populations from Trans-Omics for Precision Medicine Project. Stroke 53, 875–885.

12. UK Biobank Outcome Adjudication Group. (2022). UK Biobank Algorithmically-derived outcomes (ADOs), Version 2. In., pp 24–27.

13. Wang, K., Li, M., and Hakonarson, H. (2010). ANNOVAR: functional annotation of genetic variants from high-throughput sequencing data. Nucleic Acids Res 38, e164.

