## Supplemental materials for "Exome Array Analysis of 9,721 ischemic stroke cases from the SiGN Consortium"

Supplement

Huichun Xu[1], Kevin Nguyen[1], Brady Gaynor[1], Hua Ling[2], Wei Zhao[3,4], Patrick F. McArdle[1], Timothy O'Connor[1], O Colin Stine[5], Kathleen A. Ryan[1], Megan Lynch[1], Jennifer A. Smith[3,4], Jessica D. Faul[3], Yao Hu[6], Jeffrey W. Haessler[6], Myriam Fornage[7], Charles Kooperberg[6], the Trans-Omics for Precision Medicine (TOPMed) Stroke Working Group, James A. Perry[1], Charles C. Hong[8], John W. Cole[9], Elizabeth Pugh[2], Kimberly Doheny[2], Sharon L.R. Kardi[4], David R. Weir[3], Steven J. Kittner[9], Braxton D. Mitchell[1,10] for the SiGN Consortium

[1]Department of Medicine, University of Maryland School of Medicine, Baltimore, MD

[2]Center for Inherited Disease Research, Dept. of Genetic Medicine, Johns Hopkins University School of Medicine, Baltimore, MD

[3]Survey Research Center, Institute for Social Research, University of Michigan, Ann Arbor, MI

[4]Department of Epidemiology, School of Public Health, University of Michigan, Ann Arbor, MI

[5]Department of Epidemiology & Public Health, University of Maryland School of Medicine, Baltimore, MD

[6]Public Health Sciences Division, Fred Hutchinson Cancer Research Center, Seattle, WA

[7]Institute of Molecular Medicine, McGovern Medical School, University of Texas Health Science Center at Houston, Houston, TX

[8]Department of Cardiology Medicine, University of Maryland School of Medicine, Baltimore, MD

[9]Department of Neurology Medicine, University of Maryland School of Medicine, Baltimore, MD

[10] Geriatrics Research and Education Clinical Center, Baltimore Veterans Administration Medical Center, Baltimore, MD

Correspondence:
Dr. Mitchell;
Dr. Xu

Detailed quality control procedures

***Stage 1 QC procedures – SNP filtering (Table 2):*** Detailed quality control analyses were performed on the SiGN and HRS arrays separately to exclude duplicate SNPs, SNPs with poor QC score, high missing genotype rates, etc. An additional set of QC analyses was performed after combining the two cohorts to ensure matching of SNP names, elimination of SNPs with differential missingness, and elimination of SNPs with large AF differences between cases vs gnoMAD controls (Table 2).

Prior to current EWAS analysis, SIGN and HRS genotype data had undergone QC and cleaning at the CIDR genotyping core and then in the Data Centers of University of Washington and University of Michigan, respectively, as previously described. [1, 2] SIGN and HRS genotyping were both performed at the same core facility (CIDR) using a similar pipeline, although at different times. We first merged the exome contents of SiGN and HRS data and then implemented additional QC and filtering more specific for rare or low frequency variants as below because prior QC was implemented to address genome-wide association studies using common variants from GWAS arrays.

(1) Merge SIGN exome content and HRS exome content: SIGN exome data (4,278,837 SNPs) and HRS exome data (228,088 SNPS) were subjected to strand/allele alignment and quality check using McCarthy Group Tools (https://www.well.ox.ac.uk/~wrayner/tools/) against HRC-1000G reference genome (HRC.r1-1.GRCh37.wgs.mac5.sites.tab), and then were merged in Plink, resulting in a merged genotype data with 198811 variants and 22199 subjects.

(2) Variant filtering based on Illumina Exome array design and annotation: By design, the Illumina exome array contains content in addition to exome coding or splicing related variants, such as common variants and randomly selected synonymous SNPs included for genotyping QC and method development, and ancestry informative variants. We excluded this extra content (10413 probes) using annotation provided by the CHARGE consortium using the file SNPInfo_HumanExome-12v1_rev7.tsv (https://www.chargeconsortium.com/main/exomechip), leaving 188,398 SNPs in the merged file (variants categorized as exonic, frameshift, ncRNA_exonic, nonsynonymous, stopgain, stoploss, synonymous, cRNA_splicing, or splicing) further QC filtering.

We implemented additional QC filtering below to further ensure the quality of exome variants in the merged exome data. Specifically, we excluded SNPs (not in sequential order):

(3) with genotype AT or GC if their allele frequencies are between 40-60% (132 SNPs)

(4) if HWE p values < 1E-5 in EUR samples or < 1E-10 in AFR samples within non-stroke HRS cohort (162 SNPs); exclude SNPs if HWE p values < 1E-5 in nonstroke EUR samples within SIGN cohort (51 SNPs)

(5) with extreme Hardy-Weinberg disequilibrium if HWE p values < 1E-20 in EUR samples or < 1E-10 in AFR samples within stroke cases (3552 SNPs, 3534 of which were also excluded by other criteria)

(6) with > 0 discordant call between technical duplicate samples (n = 205 SNPs), including 29 HRS samples assayed in both SIGN Illumina 5M plus exome array and Illumina exome array, and 22 WUSTL samples assayed in both SIGN Illumina 5M plus exome array and Illumina exome array.

(7) with differential frequencies between nonstroke control samples in SIGN (EUR samples only) and nonstroke control samples in HRS (EUR samples only) based on EWAS P value< 1E-3 using SAIGE, controlling for sex and PC1-5 (n = 147 SNPs)

(8) with MAC<10 in all cases (or?) controls (34948+43777 SNPs)

(9) with missing rate in SIGN or HRS > 2.5 % (1145 SNPs)

(10) having MAF< 1% and with missing rate in SIGN or HRS > 0.008 (43289 SNPs)

(11) that are possibly under-called in SIGN by excluding those SNPs with allele count between 0 to 3 in SIGN (all ancestries included) but more than 20 in EUR samples of HRS. This was based on the observation that SNPs with problematic genotype calling cluster often are monomorphic or nearly monomorphic. We do recognize this filter may remove true positive EWAS signals.

(12) non-autosomal SNPs (3478 SNPs)

(13) that were manually reviewed and marked as technical failure in GenomeStudio clustering files of HRS cohort (n = 308 SNPs)

(14) whose allele frequencies showed significant difference (Fisher exact p value < 1E-5 for rare SNPs with MAF<1%, or < 1E-10 for low frequency or common SNPs with MAF≥1%) in EUR samples of HRS from non-finnish EUR control samples from GNOMAD V2 (21384 samples) (n = 1618 SNPs)

(15) with duplicate probes on the array but showing discordant allele frequencies in SIGN samples (Fisher exact p values < 0.05) (n = 144 SNPs)

After above filtering, 106,101 SNPs were kept for subsequent data cleaning, which was intended to minimize heterogeneity in genotype calling among different study sites within SIGN:

(16) We observed differential missingness for samples from particular study sites compared to other sites with SIGN. Therefore, we conducted comparison of missingness for each SNPs across study sites to identify and remove the potential heterogeneity in genotype calling across study sites with SIGN. Differential missingness was compared between each study (both stroke and nonstroke, if

any, samples) and the remaining studies within SIGN using Plink. 12691 SNPs were identified at differential missingness p value <5E-7 that had higher missingness in at least one study compared to the rest studies. The study samples showing higher missingness than samples from other studies were masked as missing for the corresponding SNPs (ranging from 0 to 9352 SNPs per study), so that study sample specific effect on genotyping will be minimized, reducing the potential bias toward EWA analysis.

(17) We then compared allele frequency differences across study sites within SIGN samples (only EUR samples considered) from stroke patients using SAIGE, controlling for sex and PC1-PC10. Only those SNPs with allele counts 10 or more in were considered for this QC. We identified 993 SNPs at p values < 1E-3. Each of these 993 SNPs were marked as missing for the corresponding study samples but kept intact for remaining studies within SIGN.

After removing the study-specific heterogeneity in genotyping, 106,101 SNPs were subjected for EWAS analysis using SAIGE, controlling for sex and PC1 to PC10. A predefined p value 4.7E-7 was used to determine statistical significance to accommodate Bonferroni correction (0.05/106101=4.7E-7).

***Step 2 QC procedures – re-review of genotype intensity plots:*** Genotype calling clustering plots of all associated SNPs passing predefined EWAS p values were reviewed independently by 2 co-authors (HX, BDM) to identify poor quality clustering and borderline plots were adjudicated by joint review.

References

1. NINDS Stroke Genetics Network, International Stroke Genetics Consortium. Loci associated with ischaemic stroke and its subtypes (SiGN): a genome-wide association study. Lancet Neurol 2016;15:174-184.

2. Health and Retirement Study. Exome Data (2006-2010) [online]. Available at: (https://hrs.isr.umich.edu/data-products/genetic-data/products#exome. Accessed 7/2/2022.

Table S1: Cases (n = 9,721) and controls (n = 12,345) by site and ancestry

| Genetic Ancestry | EUR | | | | AFR | | | | Others | | |
|---|---|---|---|---|---|---|---|---|---|---|---|
| **Study** | **con** | **case** | **EURtotal** | | **con** | **case** | **AFRtotal** | | **con** | **case** | **Other_total** |
| totalN | 9,911 | 7,921 | 17,832 | | 1,353 | 1,044 | 2,397 | | 1,081 | 756 | 1,837 |
| BASICMAR | 0 | 918 | 918 | | | | | | | 12 | 12 |
| BRAINS | 0 | 108 | 108 | | | | | | | 6 | 6 |
| GASROS | 0 | 440 | 440 | | 0 | 12 | 12 | | | 18 | 18 |
| GCNKSS | 0 | 379 | 379 | | 0 | 119 | 119 | | | 1 | 1 |
| GOTEBURG | 0 | 791 | 791 | | | | | | | 9 | 9 |
| GRAZ | 29 | 639 | 668 | | | | | | | | |
| HRS | 8,613 | 0 | 8,613 | | 1,348 | 0 | 1,348 | | 1,081 | 0 | 1,081 |
| ISGS | 0 | 66 | 66 | | 0 | 120 | 120 | | | 1 | 1 |
| KRAKOW | 776 | 951 | 1,727 | | | | | | | 1 | 1 |
| LEUVEN | 468 | 482 | 950 | | | | | | | | |
| LUND | 0 | 645 | 645 | | | | | | | 6 | 6 |
| MCISS | 0 | 504 | 504 | | 0 | 90 | 90 | | | 36 | 36 |
| MIAMISR | 0 | 138 | 138 | | 0 | 101 | 101 | | | 60 | 60 |
| NHS | 0 | 316 | 316 | | | | | | | | |
| NOMAS | 0 | 115 | 115 | | 0 | 108 | 108 | | | 140 | 140 |
| OAI | 25 | 0 | 25 | | 5 | 0 | 5 | | | | |
| REGARDS | 0 | 175 | 175 | | 0 | 131 | 131 | | | 5 | 5 |
| SPS3 | 0 | 362 | 362 | | 0 | 144 | 144 | | | 454 | 454 |
| SWISS | 0 | 202 | 202 | | 0 | 1 | 1 | | | 2 | 2 |
| WHI | 0 | 420 | 420 | | 0 | 34 | 34 | | | 4 | 4 |
| WUSTL | 0 | 270 | 270 | | 0 | 184 | 184 | | | 1 | 1 |

### Genotyping intensity plots for 15 exome-wide significant SNPs

| SNP | rs number | Gene |
| --- | --- | --- |
| exm1345082 | rs192153785 | GH2 |
| exm1501517 | rs140922537 | ZNF765 |
| exm1562153 | rs143510517 | TPTE |
| exm21949 | rs373898350 | NBPF1 |
| exm365204 | rs141845742 | SPATA16 |
| exm384695 | rs149905649 | DOK7 |
| exm552854 | rs62619974 | MEP1A |
| exm558342 | rs199585353 | PRIM2 |
| exm615057 | rs375144101 | TRGC1 |
| exm791656 | rs142792732 | DDX31 |
| exm90767 | rs372423248 | SEC22B |
| exm90783 | rs373433490 | SEC22B |
| exm913753 | rs79336999 | PATL1 |
| exm-rs507666 | rs507666 | ABO |
| exm-rs635634 | rs635634 | ABO |

### exm1345082

#### SiGN (mostly stroke cases)

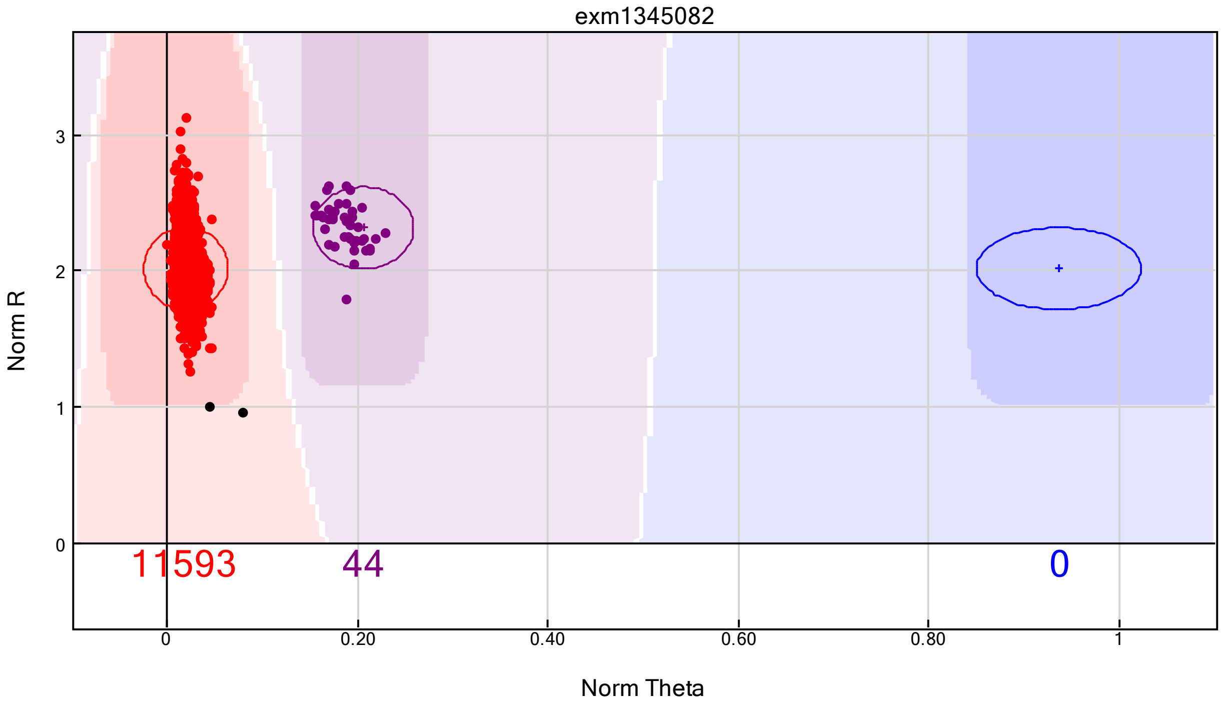


#### HRS (all non-stroke controls)

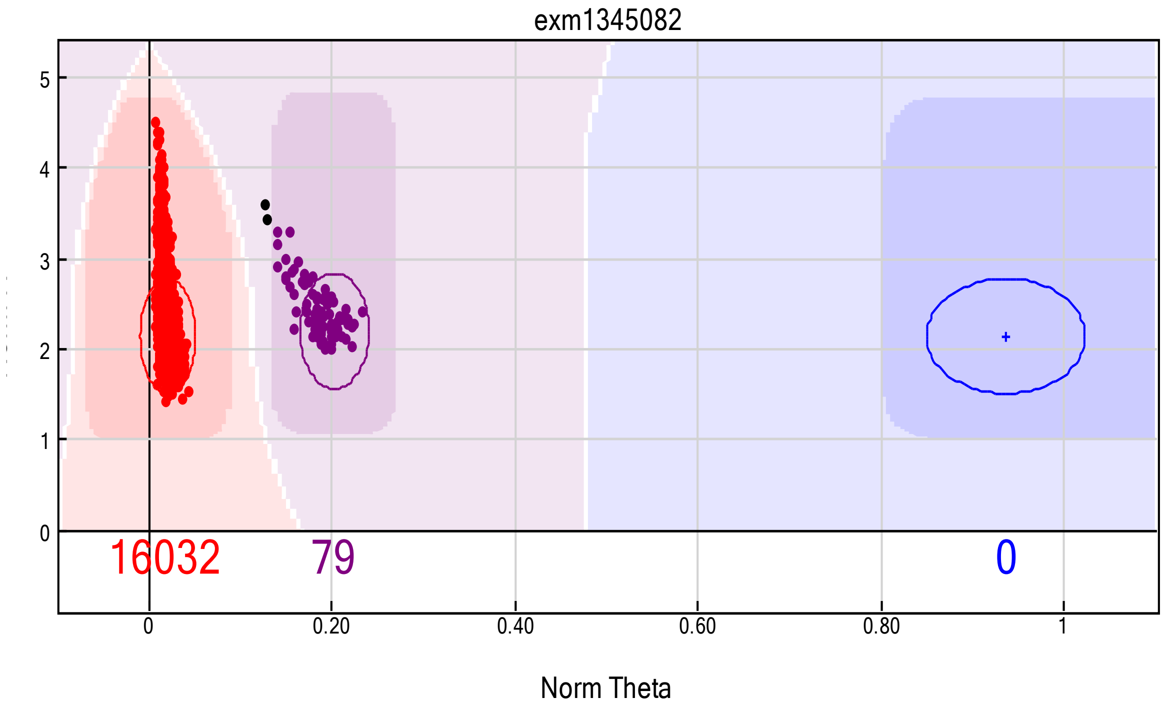

### exm1501517

#### SiGN (mostly stroke cases)

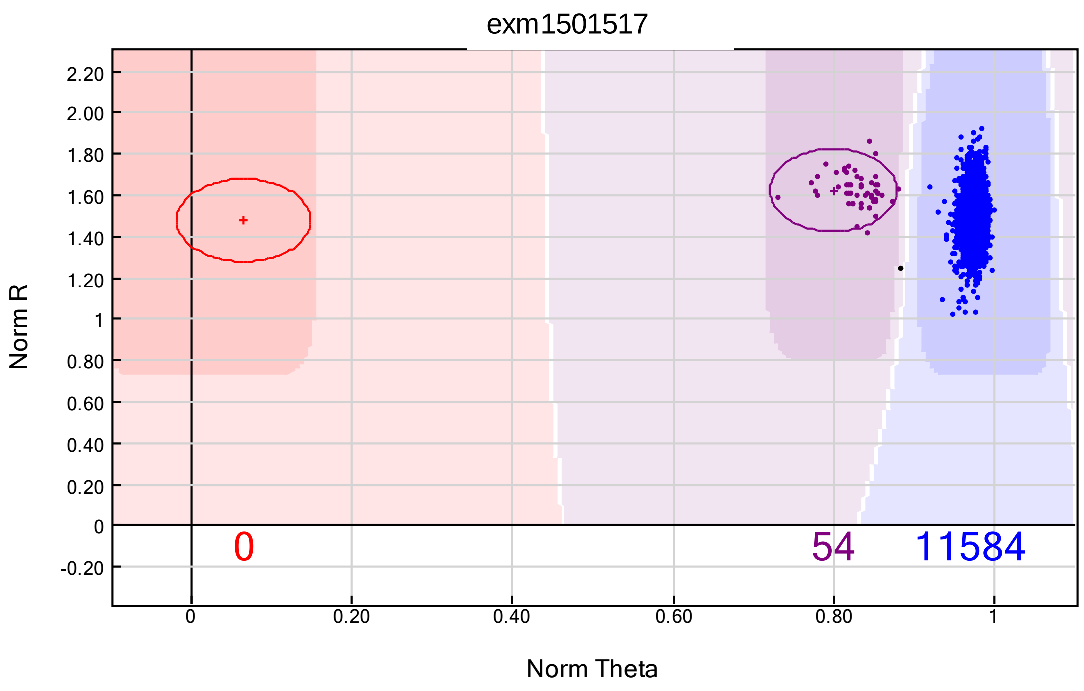


#### HRS (all non-stroke controls)

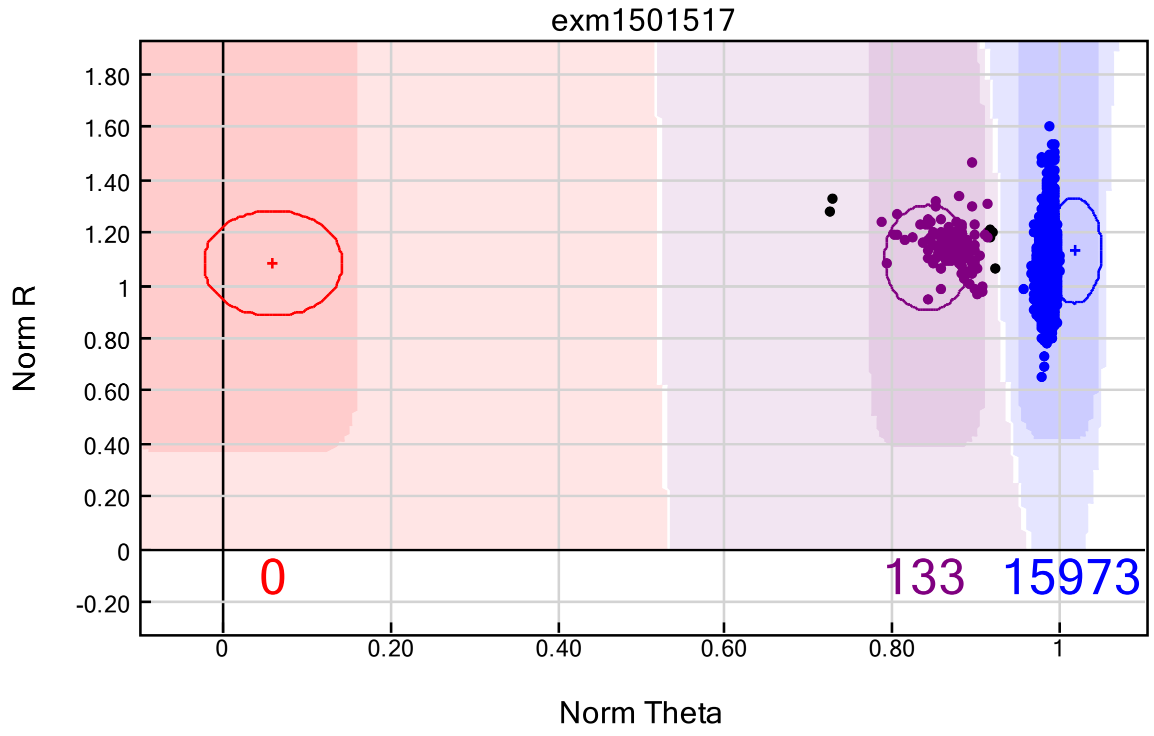

### exm1562153

#### SiGN (mostly stroke cases)

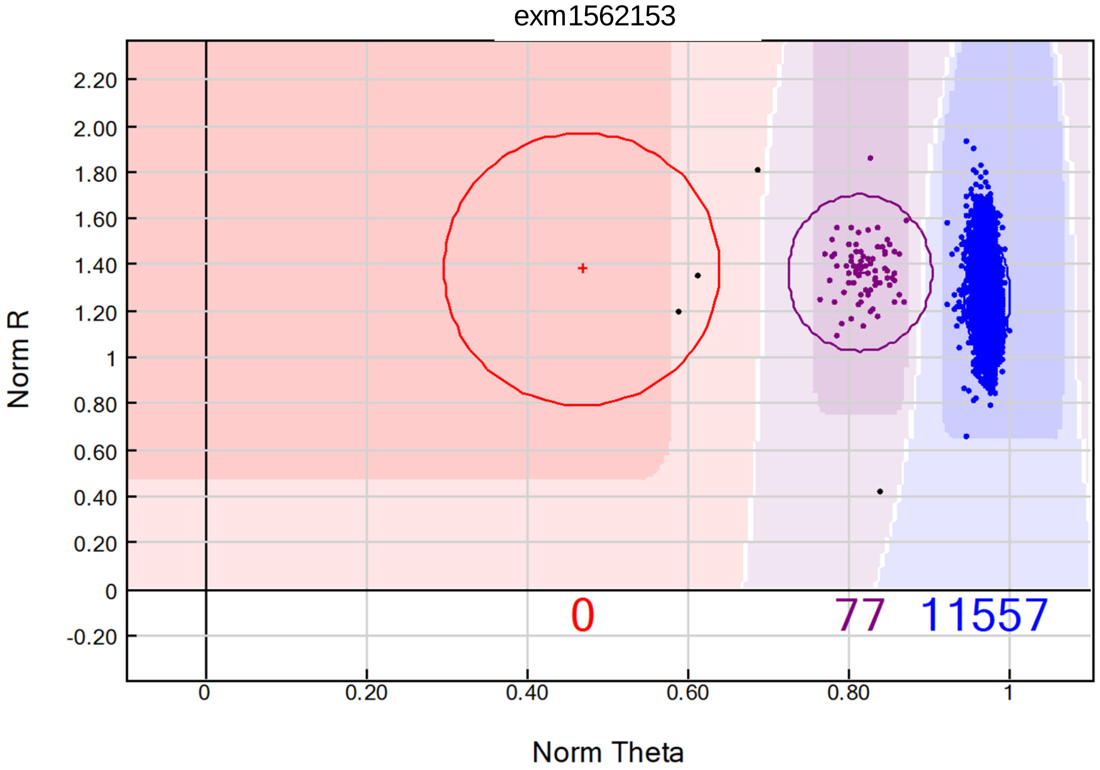


#### HRS (all non-stroke controls)

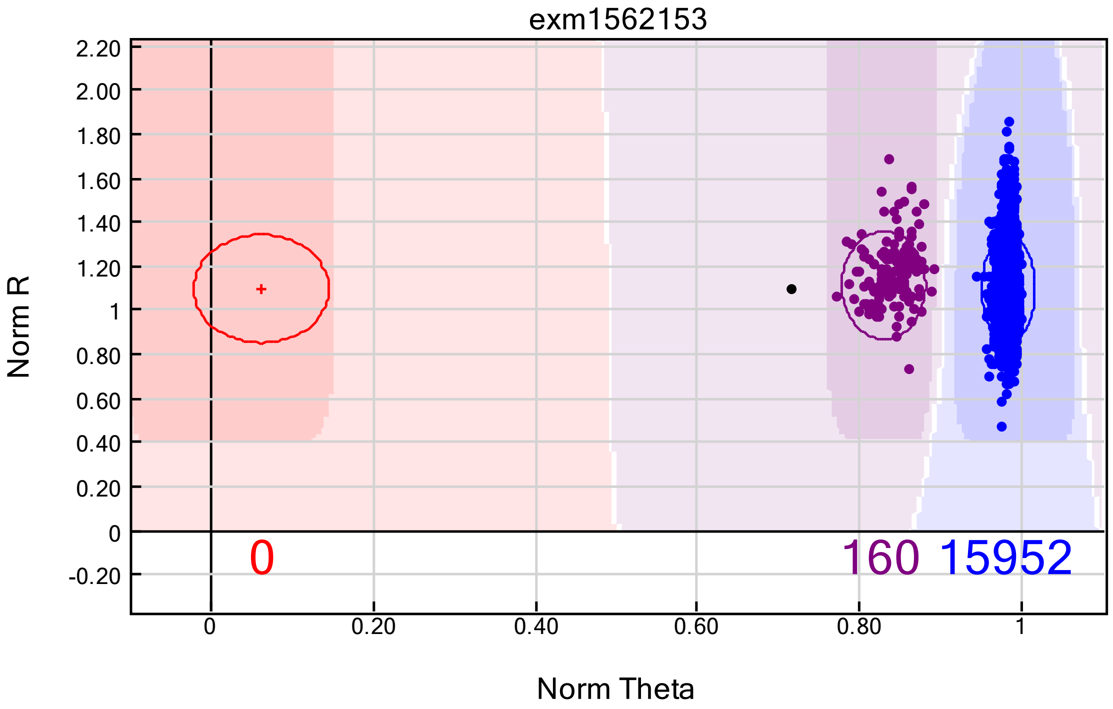

### exm21949

#### SiGN (mostly stroke cases)

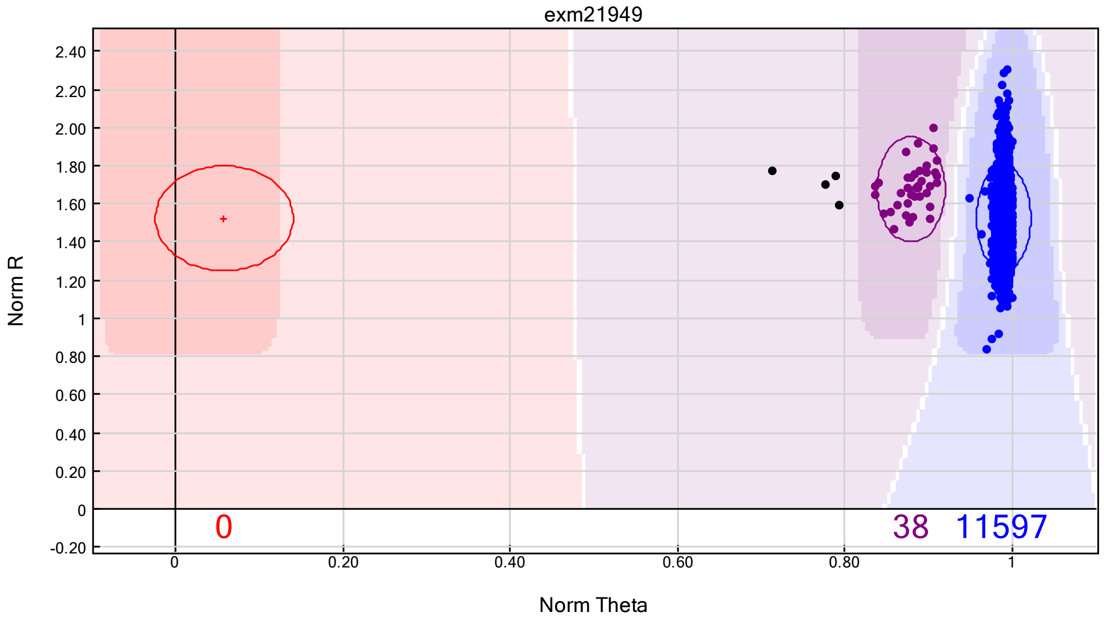


#### HRS (all non-stroke controls)

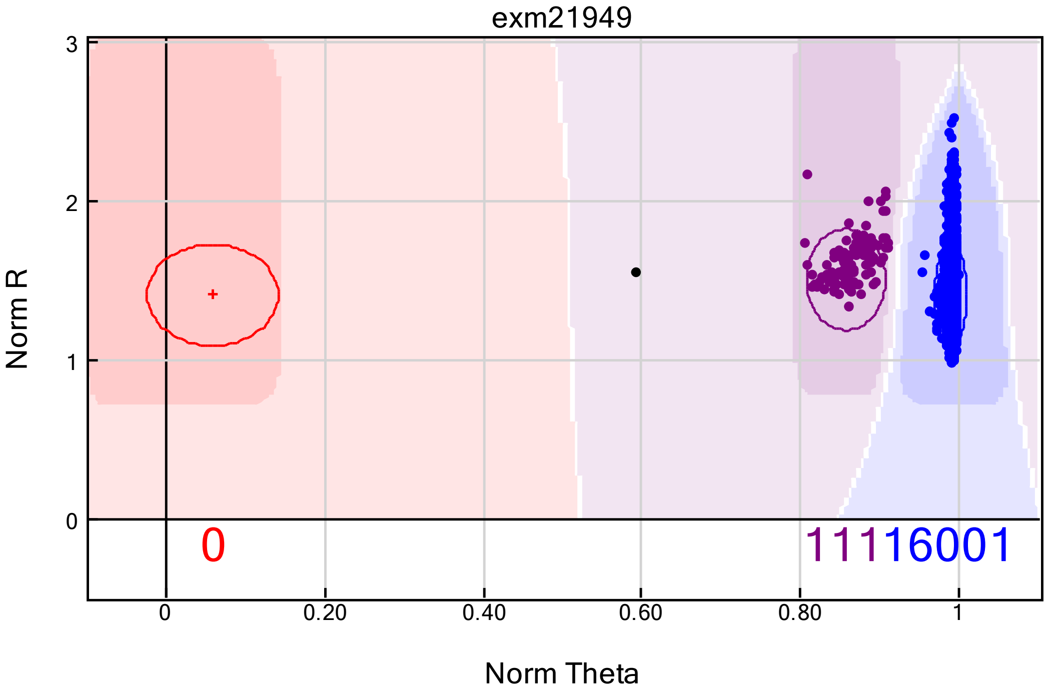

### exm365204

#### SiGN (mostly stroke cases)

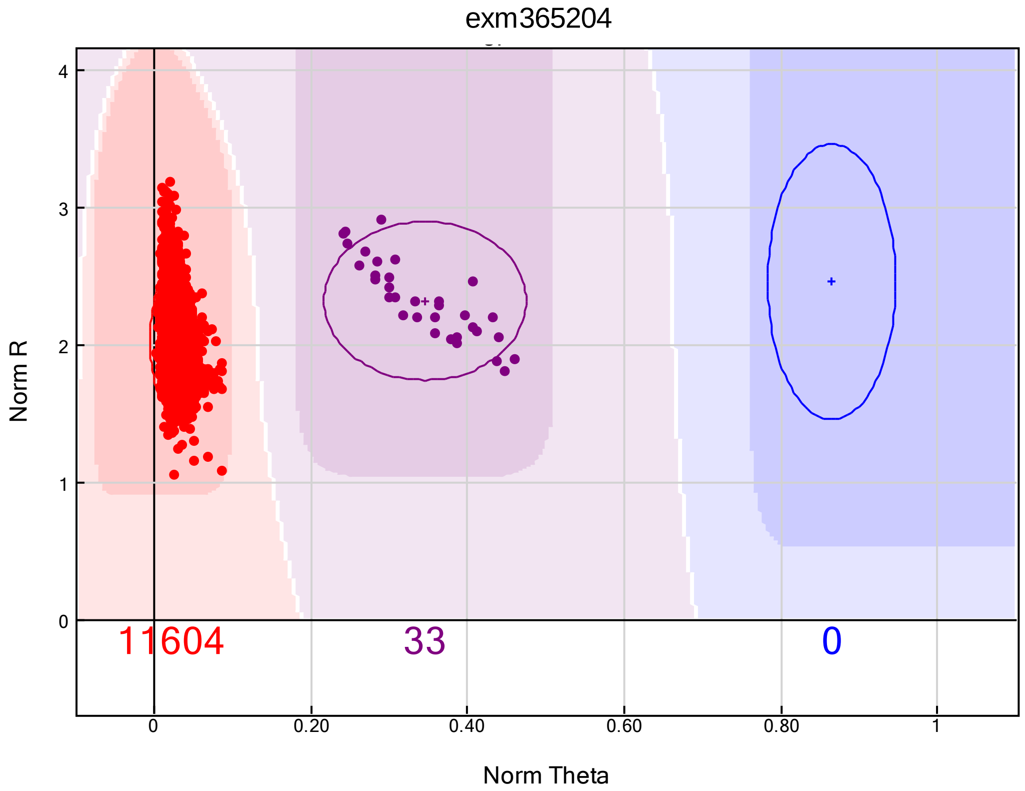


#### HRS (all non-stroke controls)

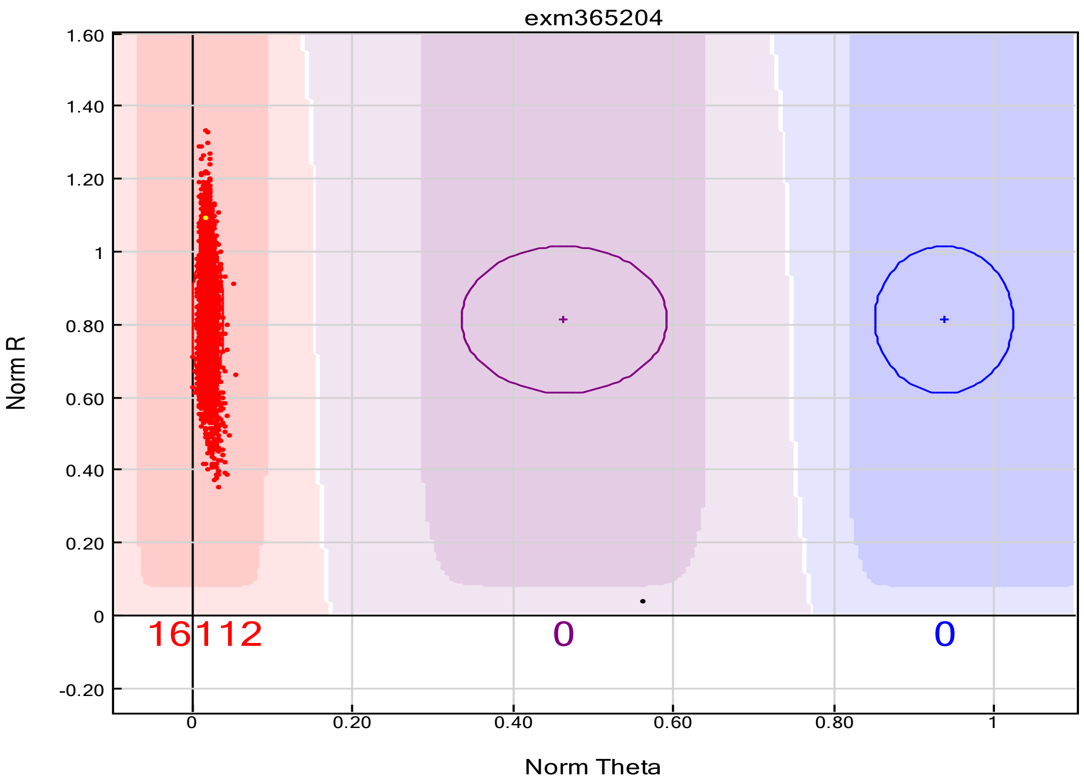

### exm384695

#### SiGN (mostly stroke cases)

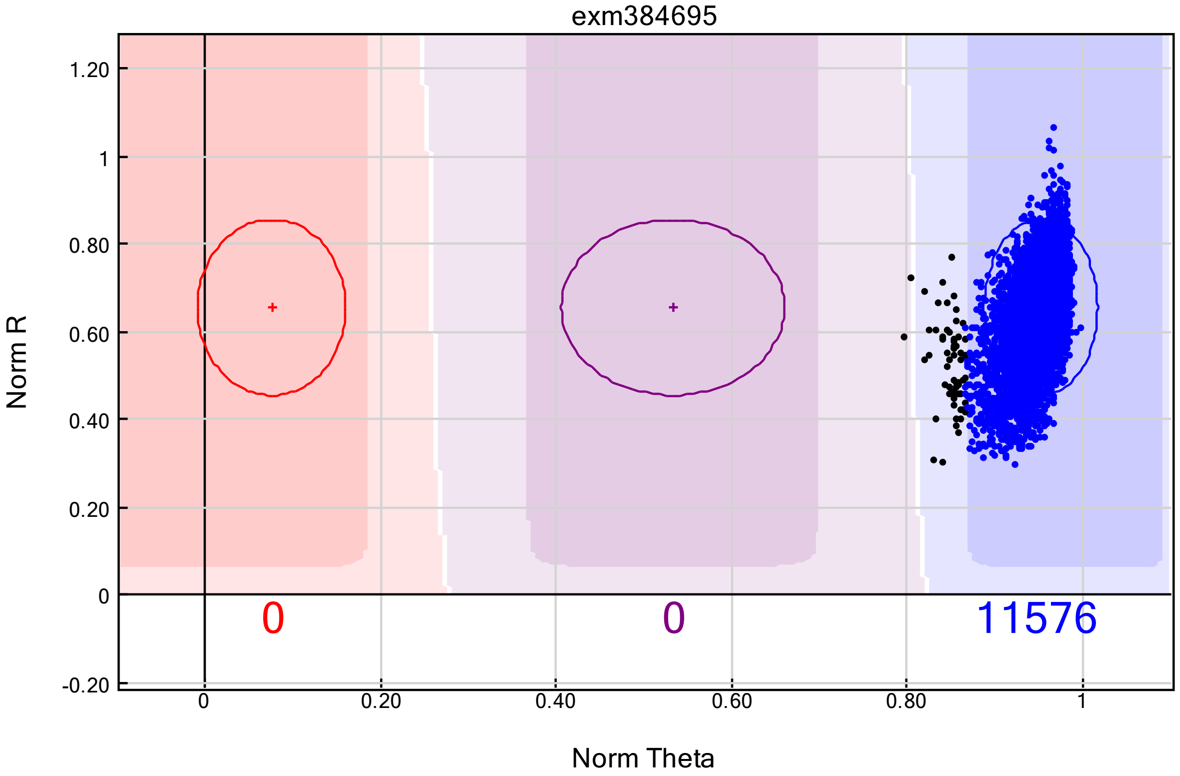


#### HRS (all non-stroke controls)

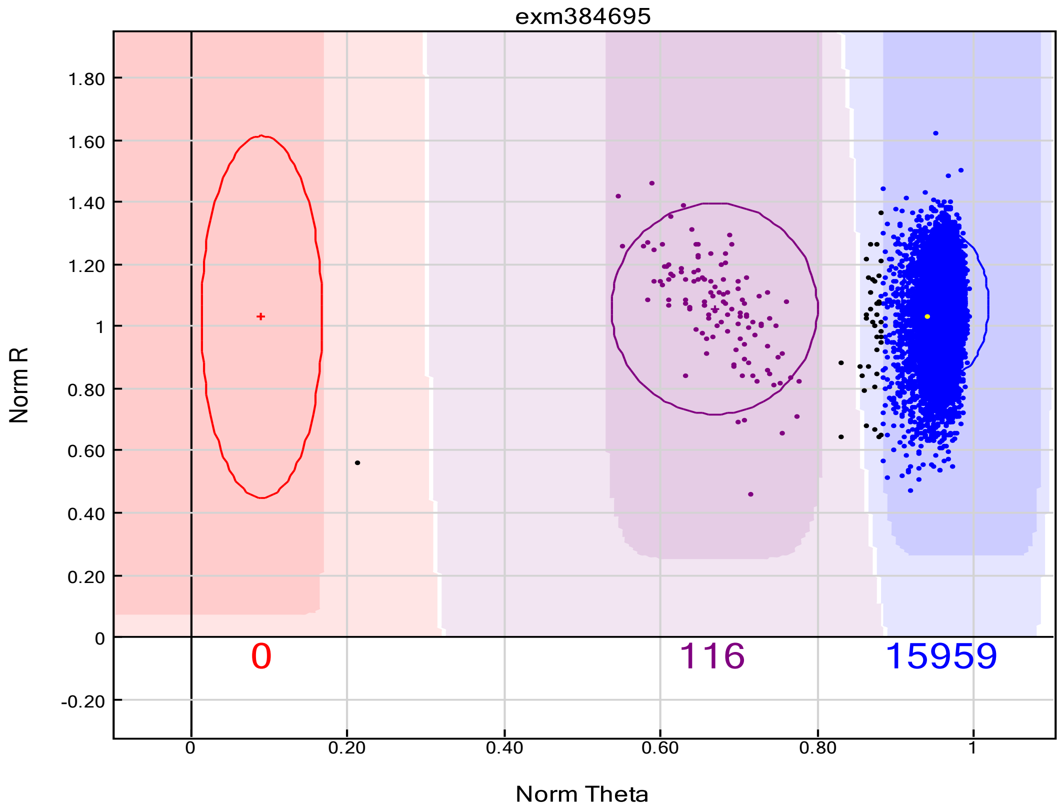

### exm552854

#### SiGN (mostly stroke cases)

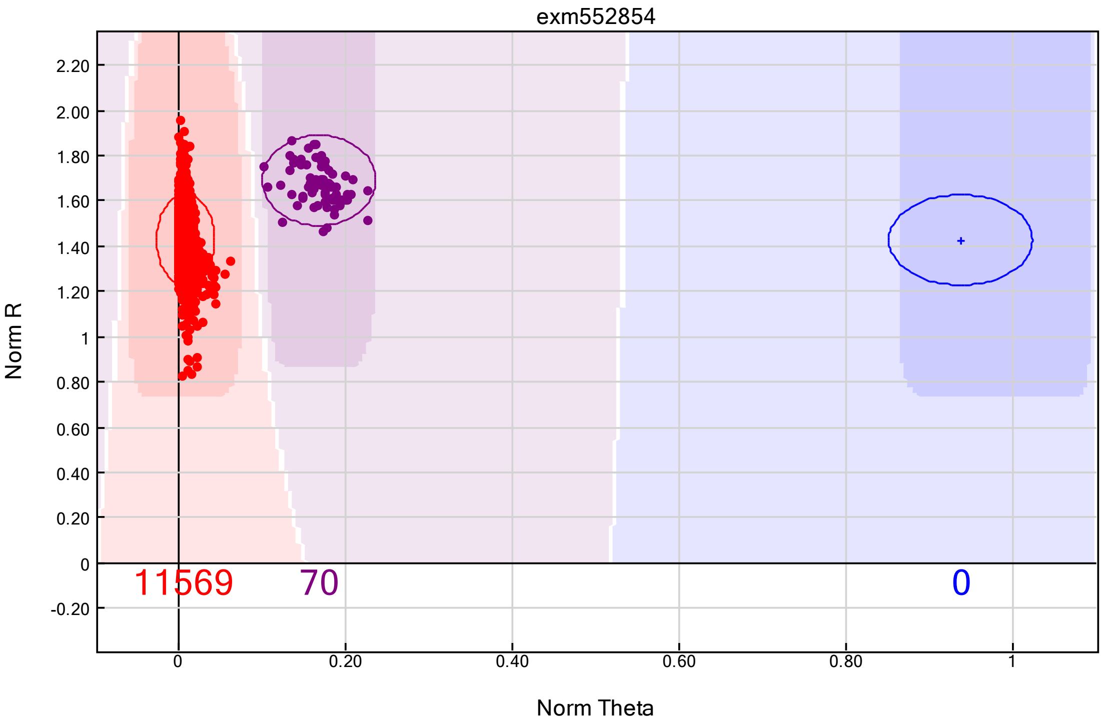


#### HRS (all non-stroke controls)

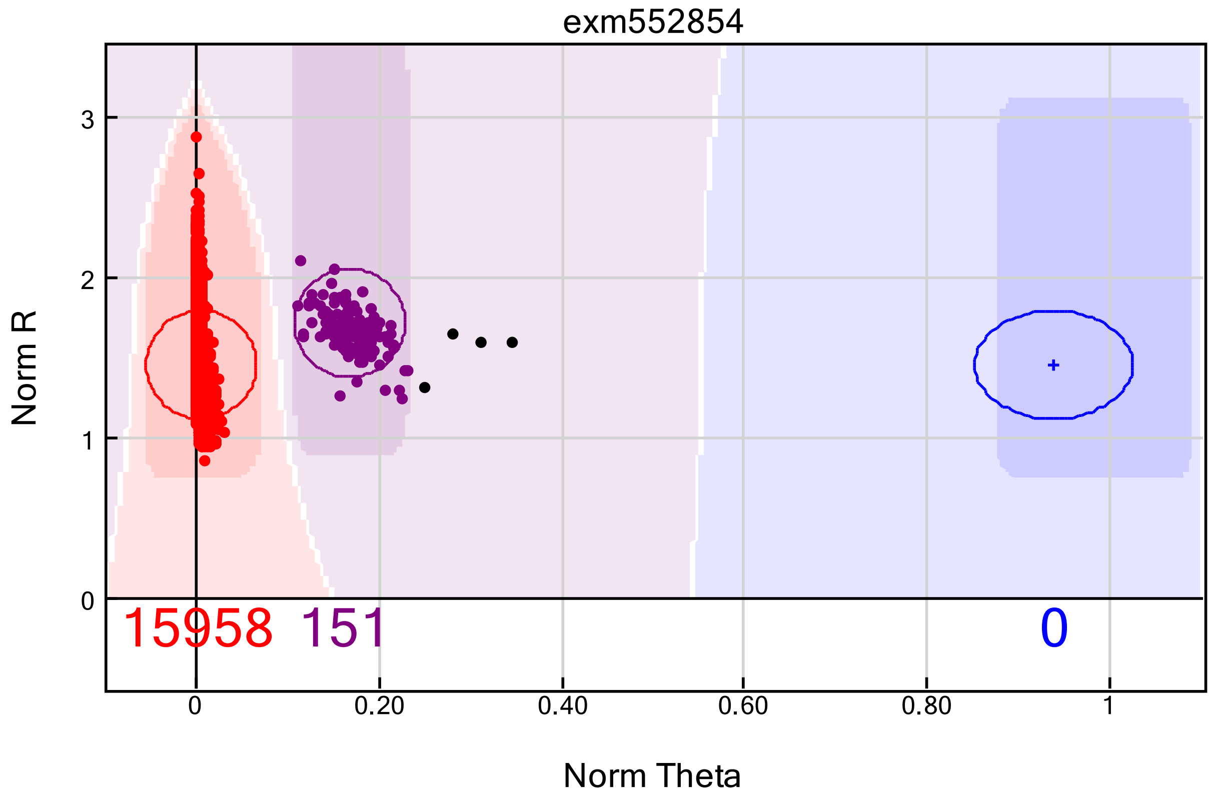

### exm558342

#### SiGN (mostly stroke cases)

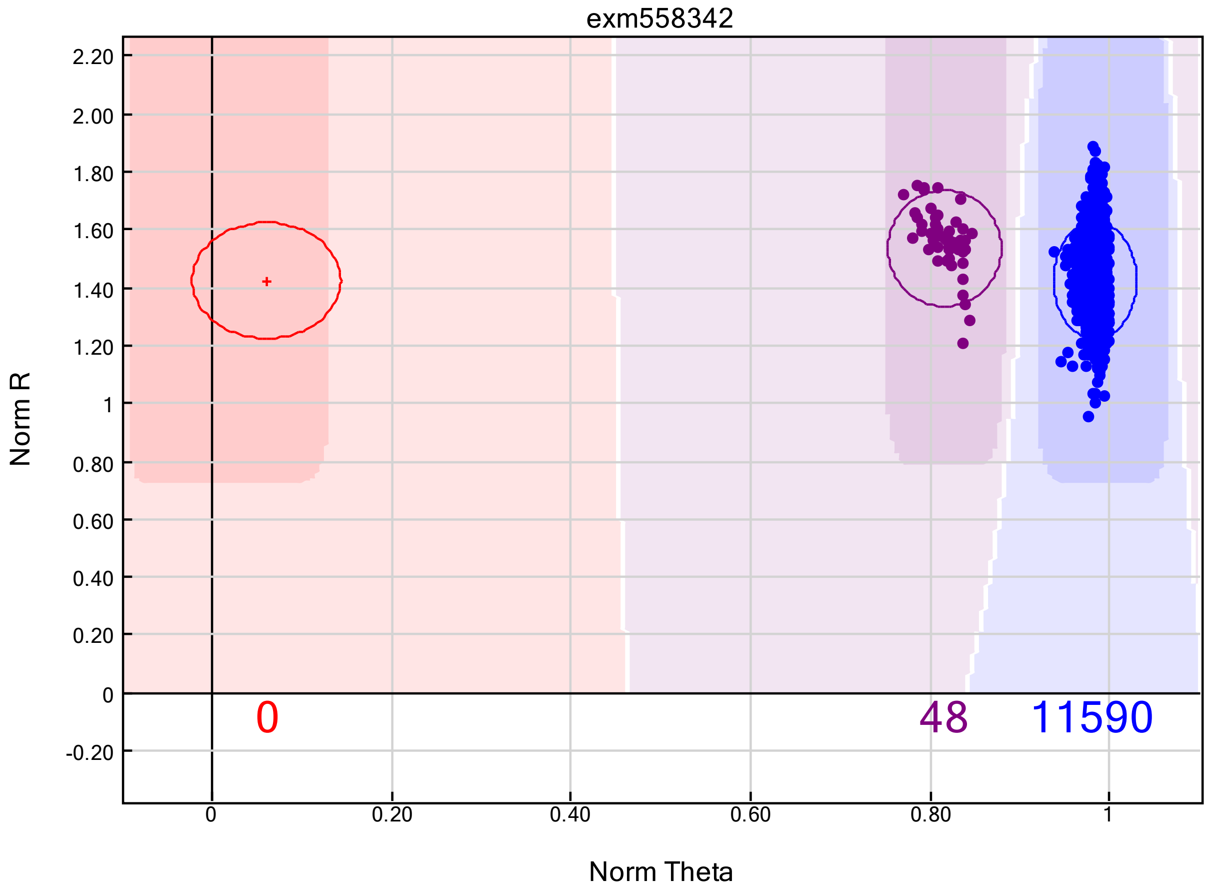


#### HRS (all non-stroke controls)

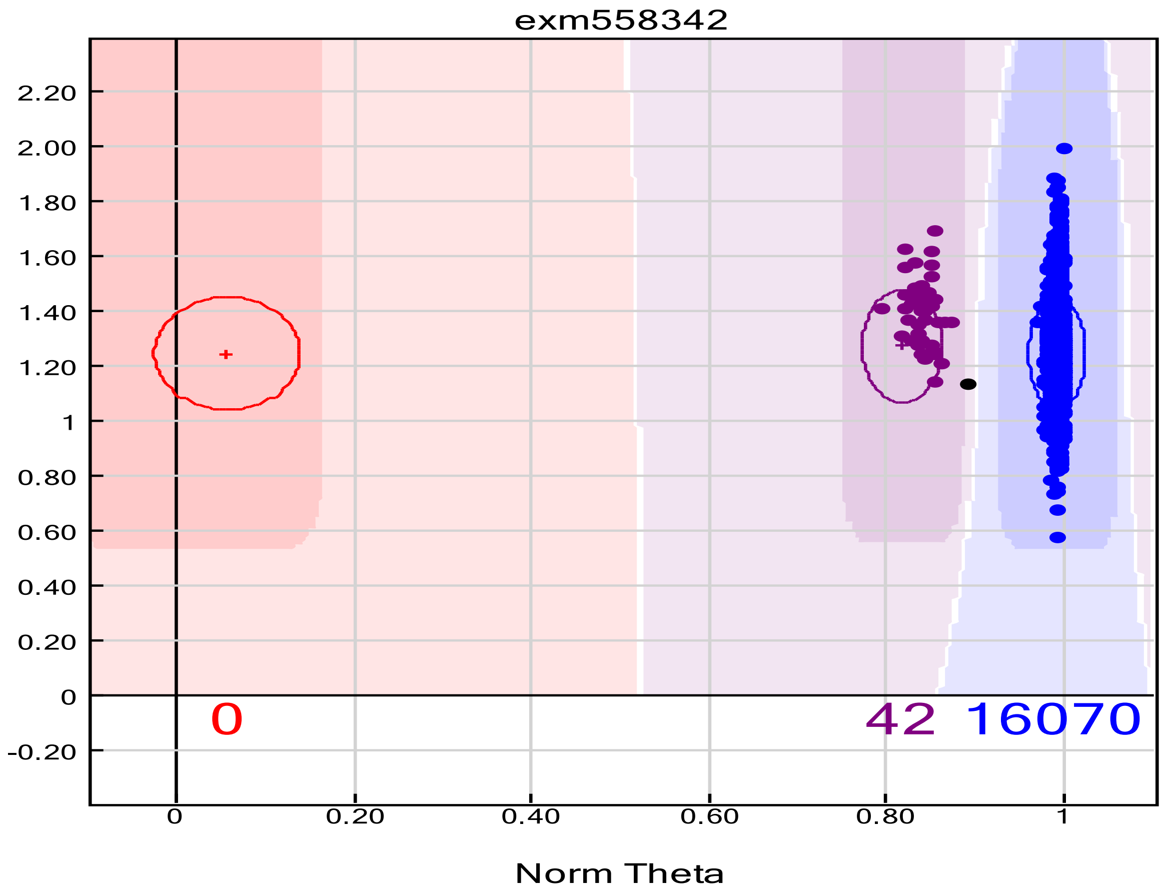

### exm615057

#### SiGN (mostly stroke cases)

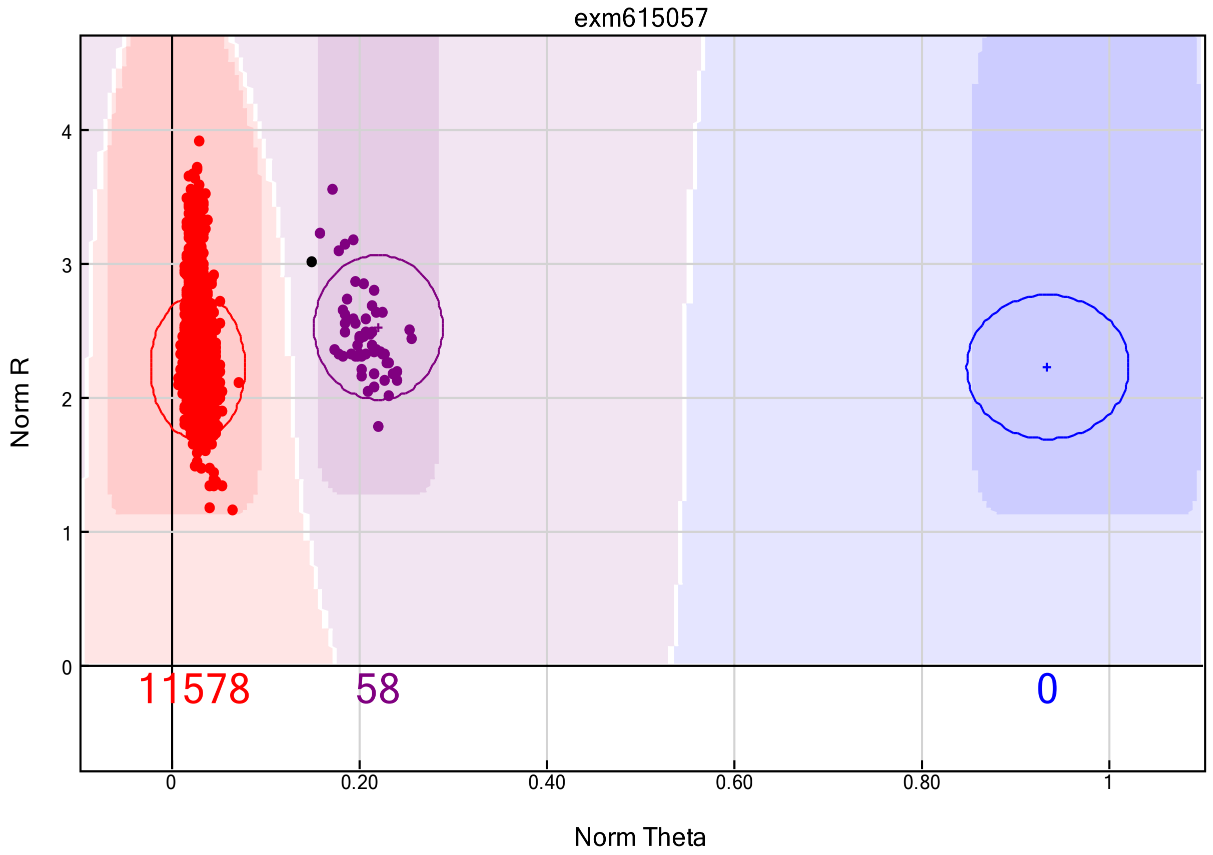


#### HRS (all non-stroke controls)

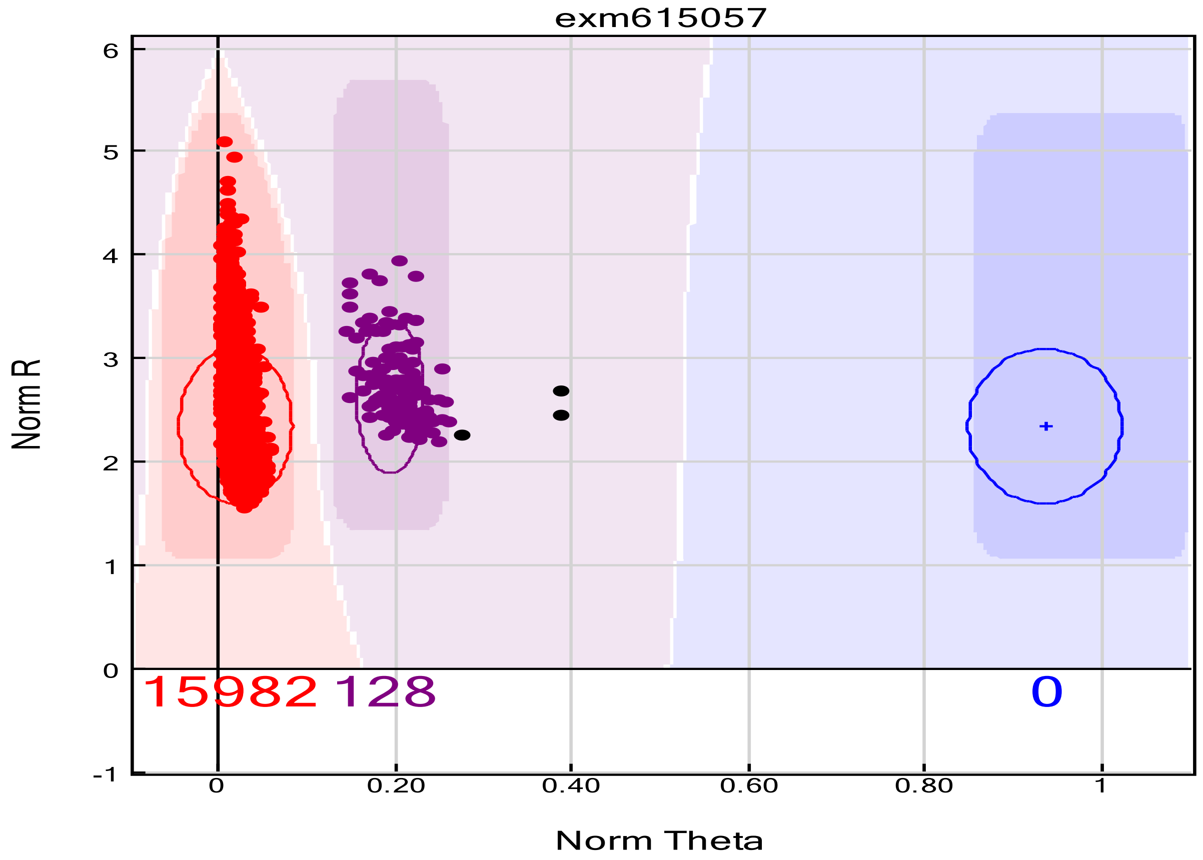

### exm791656

#### SiGN (mostly stroke cases)

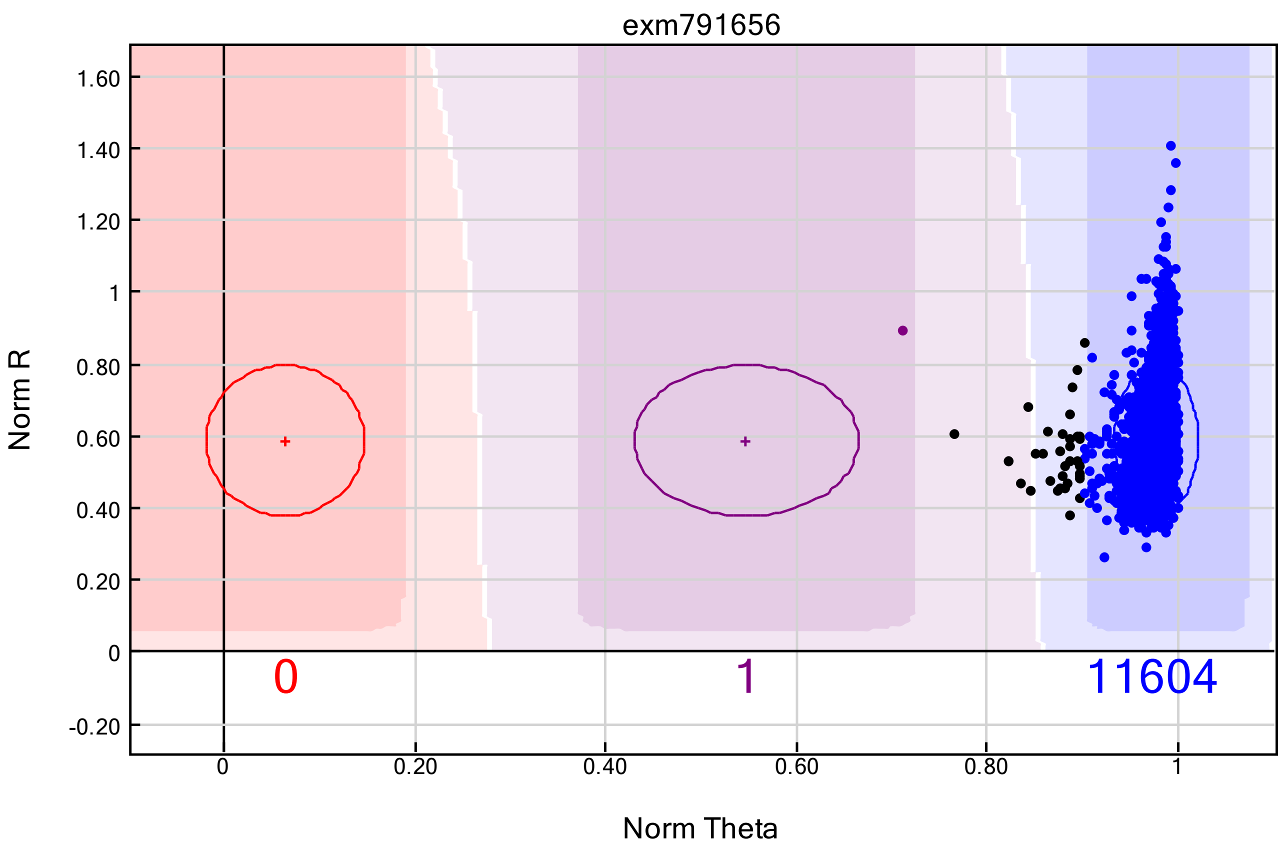


#### HRS (all non-stroke controls)

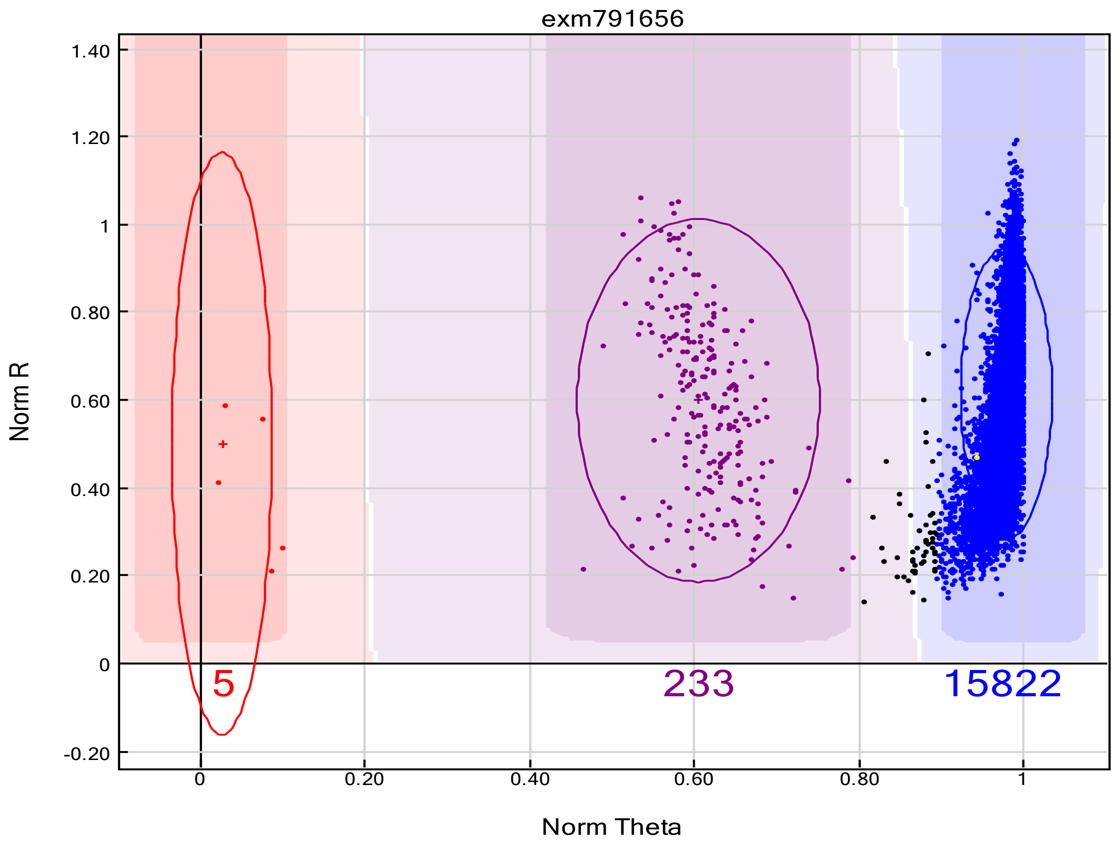

### exm90767

#### SiGN (mostly stroke cases)

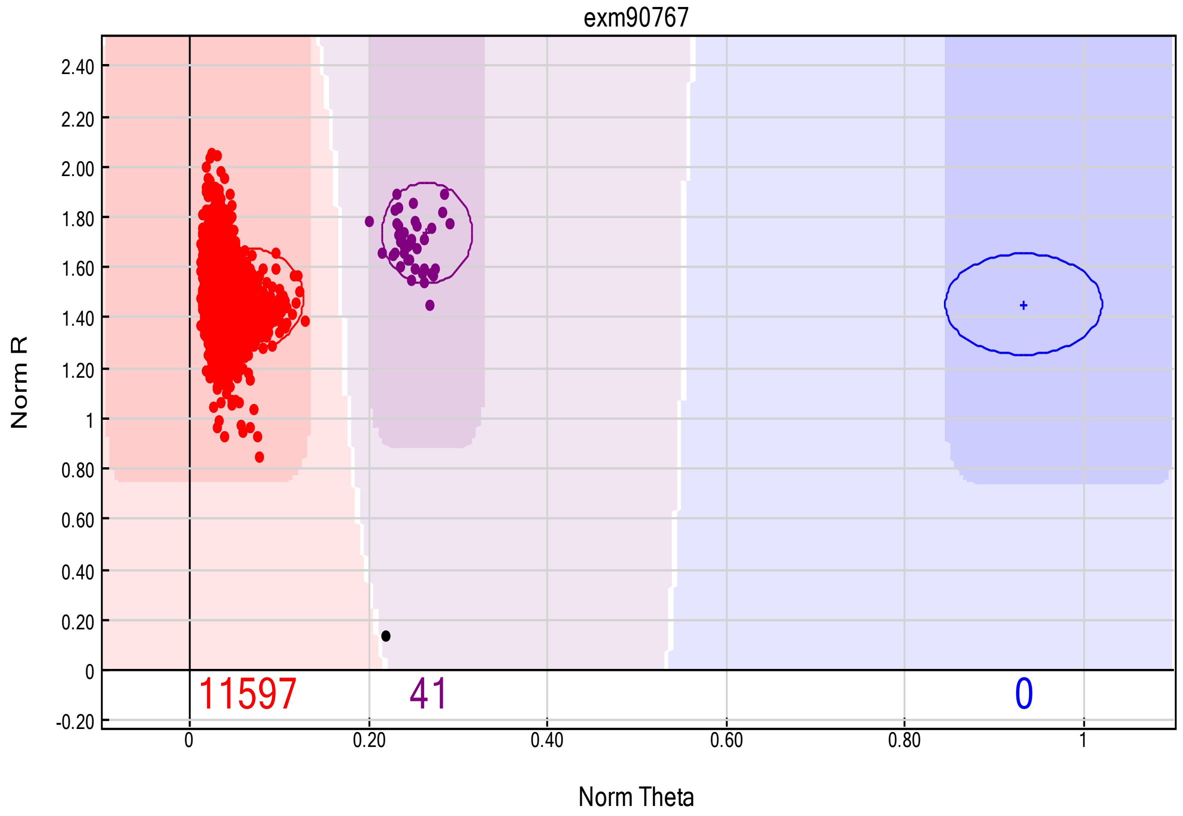


#### HRS (all non-stroke controls)

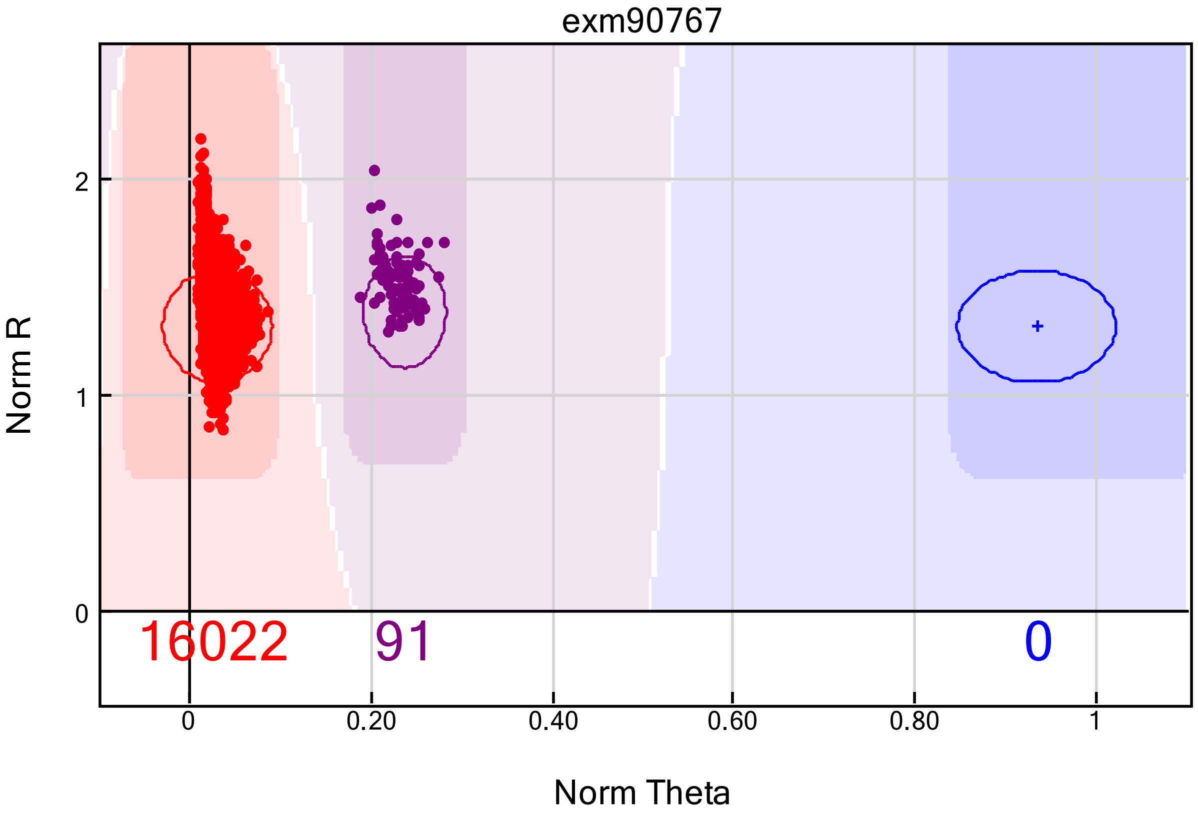

### exm90783

#### SiGN (mostly stroke cases)

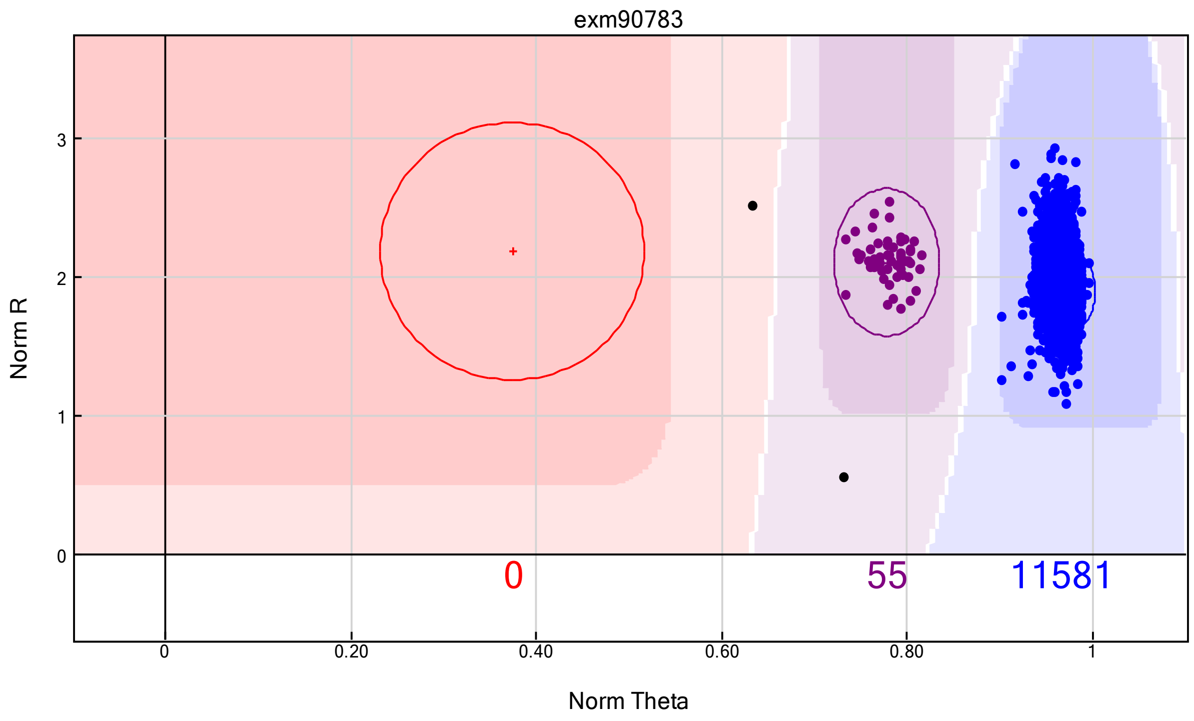


#### HRS (all non-stroke controls)

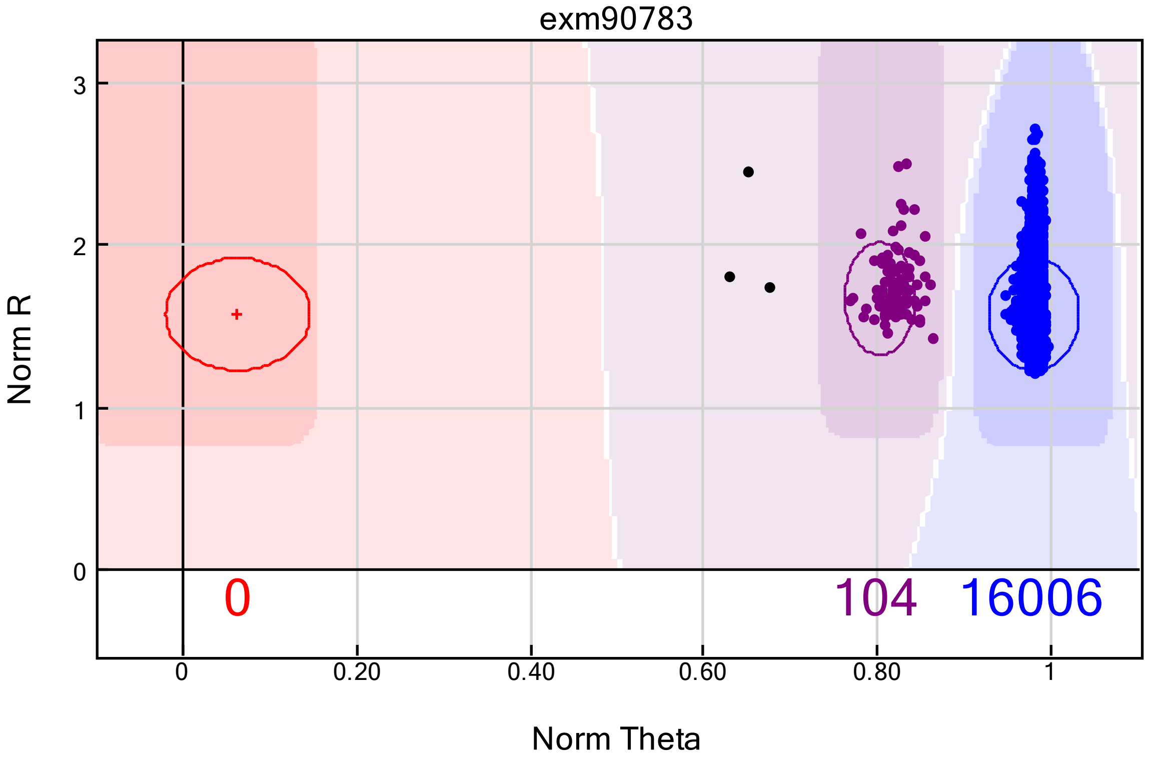

### exm913753

#### SiGN (mostly stroke cases)

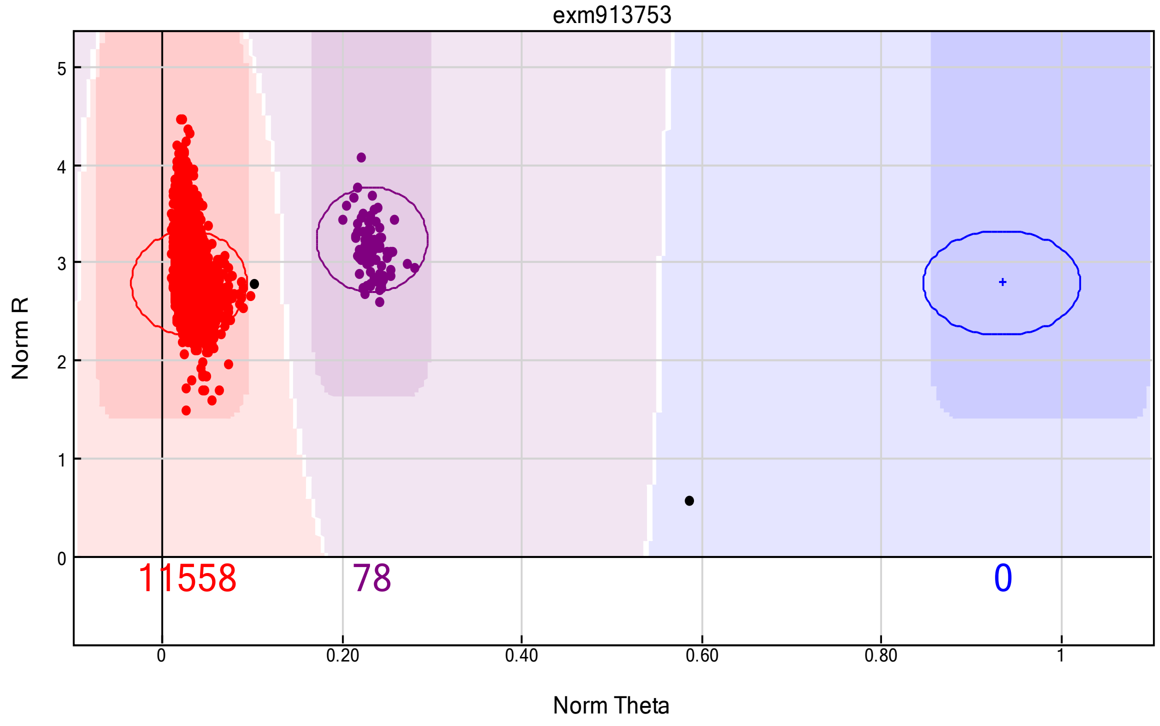


#### HRS (all non-stroke controls)

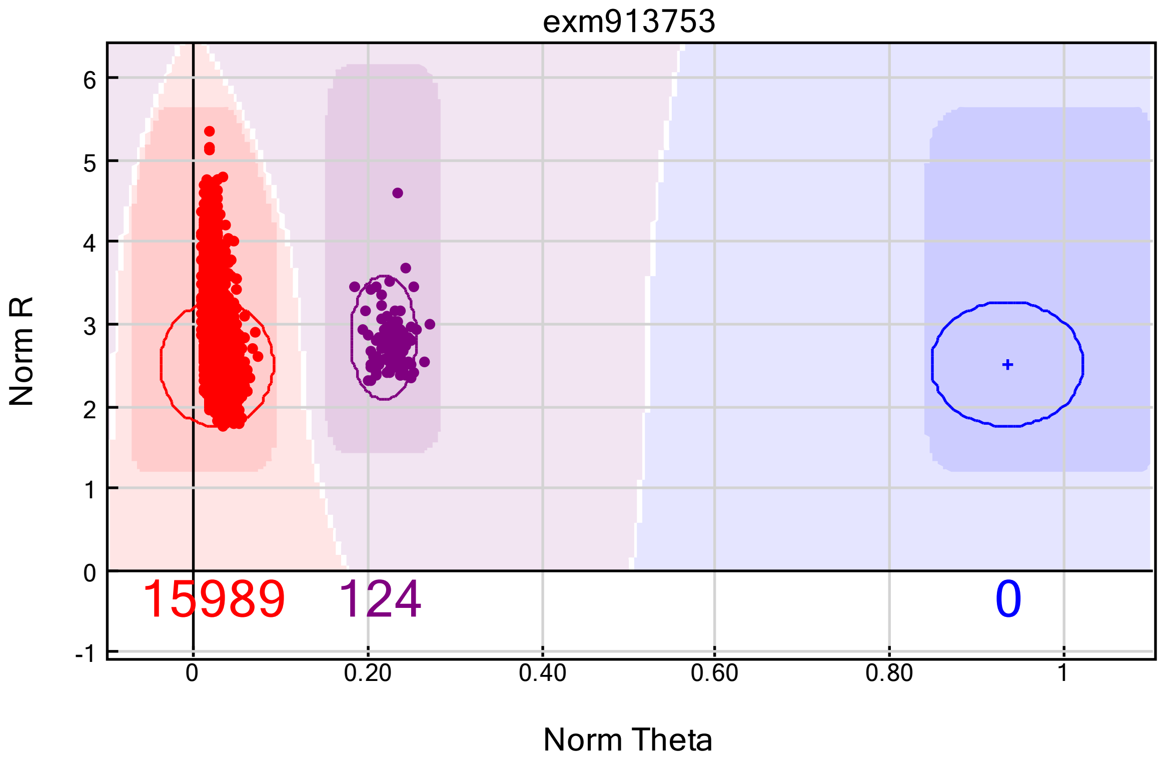

### exm-rs507666

#### SiGN (mostly stroke cases)

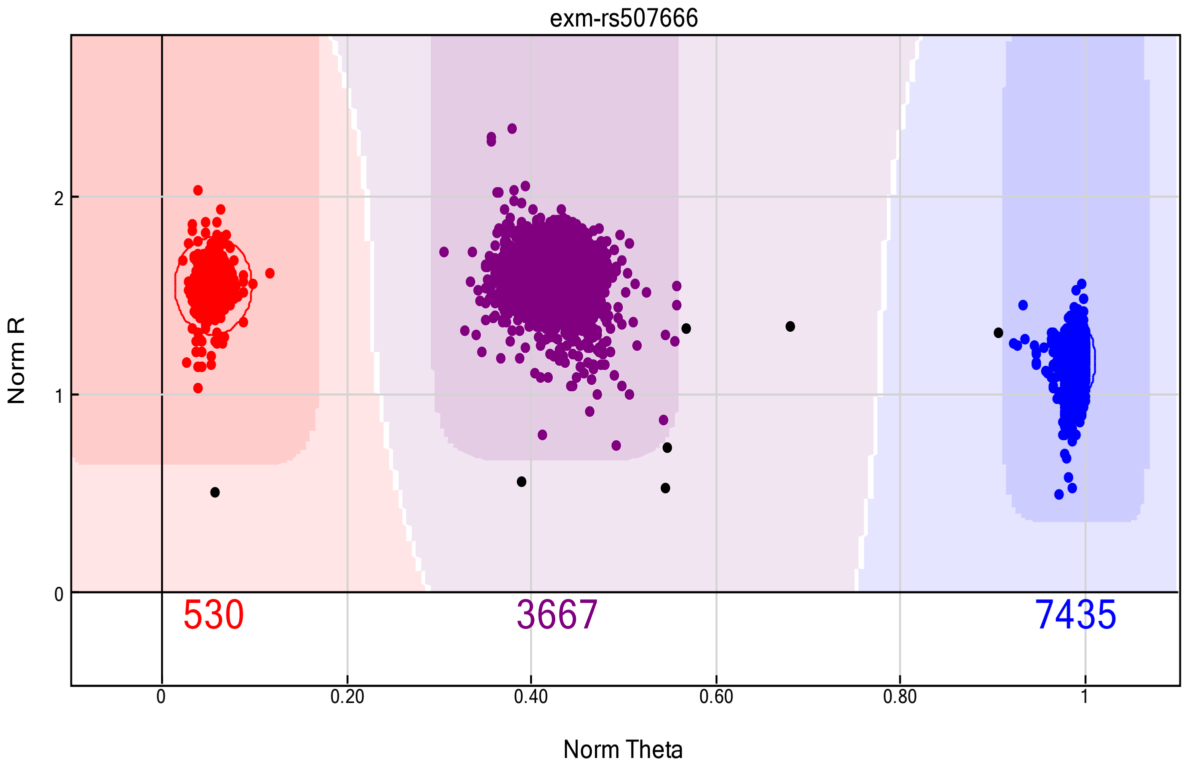


#### HRS (all non-stroke controls)

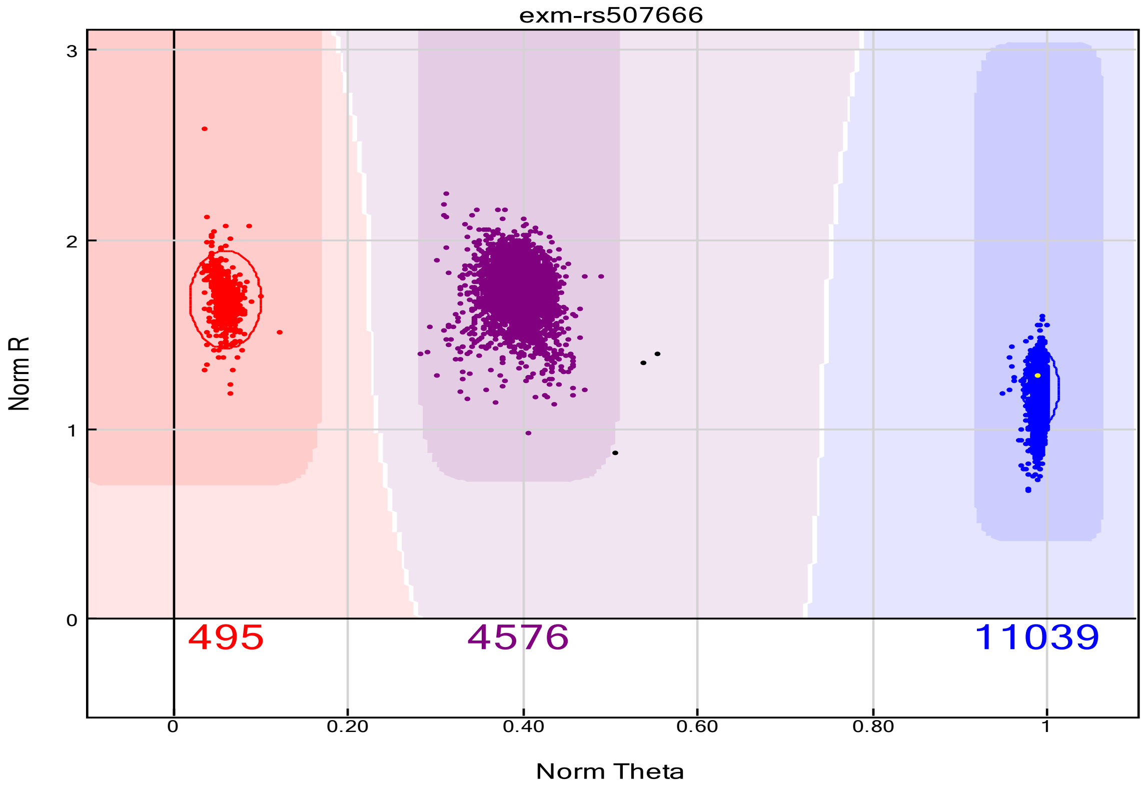

### exm-rs635634

#### SiGN (mostly stroke cases)

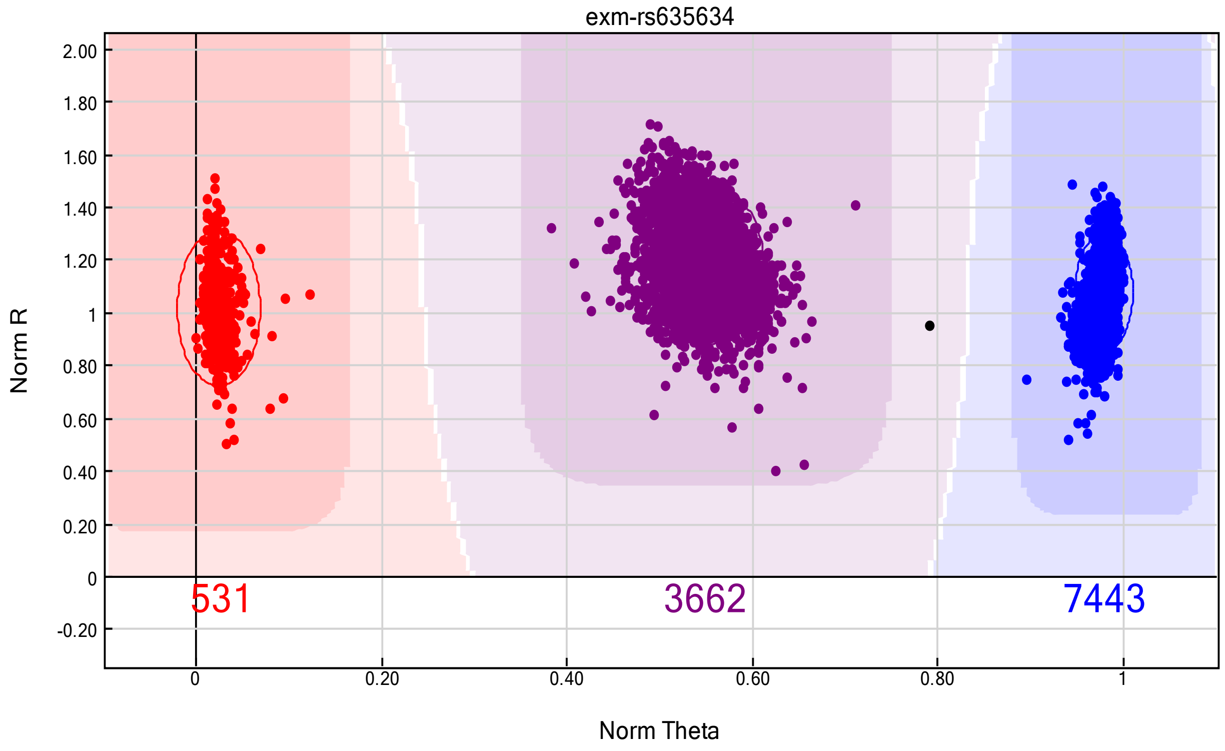


#### HRS (all non-stroke controls)

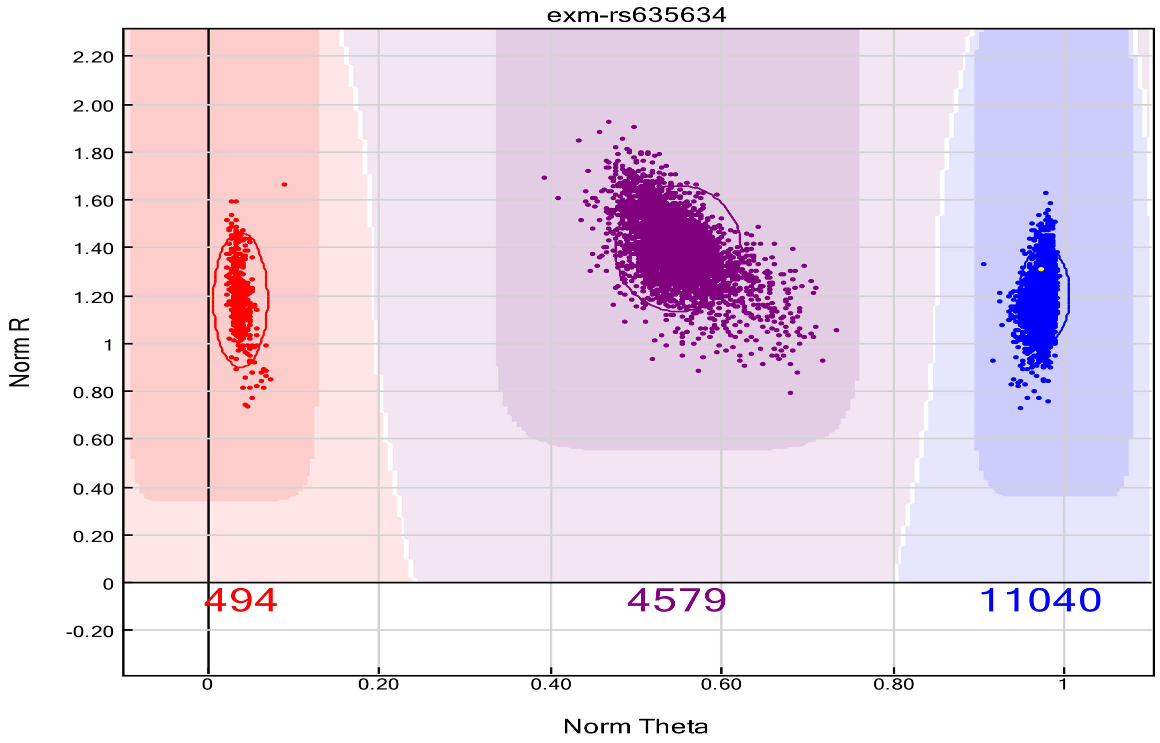
